# Does sleep impact mobility in adults? A scoping review

**DOI:** 10.1101/2025.05.05.25326996

**Authors:** Catherine Siengsukon, Mahya Beheshti, Sarah J. Donkers, Silvana L. Costa, Prasanna Vaduvathiriyan, Allison Glaser, Garrett Baber, Joy Williams

**Affiliations:** Department of Physical Therapy, Rehabilitation Science, and Athletic Training, University of Kansas Medical Center, Kansas City, KS, USA; Department of Physical Medicine and Rehabilitation, NYU Grossman School of Medicine, New York, NY, USA; School of Rehabilitation Science, University of Saskatchewan, Saskatoon, Canada; Center for Neuropsychology and Neuroscience Research, Kessler Foundation, East Hanover, NJ; Department of Physical Medicine and Rehabilitation, Rutgers –New Jersey Medical School, Newark, NJ, USA; A.R. Dykes Library, University of Kansas Medical Center, Kansas City, KS, USA; Department of Psychology, University of Kansas, Lawrence, Kansas, USA; Department of Physical Therapy, University of the Pacific, Stockton, CA, USA

## Abstract

**Objective:** To review the literature on if sleep impacts mobility in adults.

**Review method used:** This scoping review followed the Preferred Reporting Items for Systematic Reviews and Meta-Analyses extension for scoping reviews (PRISMA – ScR) guidelines and the Joanna Briggs Institute’s (JBI) updated methodology for scoping reviews.

**Data sources:** Ovid Medline, Web of Science, Embase.com, and CINAHL databases were searched. Google Scholar and hand-searching were further reviewed for grey literature.

**Review methods:** Seven of the authors participated in the data screening and extraction process. Citations were randomly divided so that each was screened separately by two team members. Similarly, data were extracted from included articles independently by two reviewers with a third dedicated to resolving discrepancies.

**Results:** The search strategy generated 8,772 references, 697 articles underwent full-text screening, and 108 eligible articles were included in the scoping review. Most studies (n = 69; 64%) included only self-reported sleep, and “walking” was the most common mobility category (n = 99). Most studies (n = 60; 56%) reported a positive association between sleep and mobility, indicating better sleep was associated with better mobility or worse sleep was associated with worse mobility. Most studies including people with cardiovascular, kidney, metabolic, mental health, neurological, and pulmonary conditions reported a positive association between sleep and mobility.

**Conclusion:** Most studies reported a positive association between sleep and mobility. However, due to the variety of sleep and mobility outcomes used, it was challenging to compare studies and synthesize results. Further, due to a relatively small sample size and variety of health conditions, conclusions cannot be drawn, and further research is needed.

## INTRODUCTION

Sleep disturbances and insufficient sleep are common issues. More than 30% of adults get less than the recommended 7 hours of sleep each night, and 50% report not feeling well-rested^1,2^ suggesting inadequate sleep quality. Insufficient sleep is particularly pronounced in older adults, with approximately 75% of older adults reporting symptoms of insomnia.^1^

Inadequate sleep and sleep disturbances have negative consequences on health and well-being, including reducing physical and cognitive function, negatively impacting participation in physical activity, and increasing risk of falls.^3–5^ Poor sleep disrupts the body’s natural restorative processes, which are crucial for muscle repair, memory consolidation, and overall recovery.^4,6^ Particularly regarding functional ability, older adults who have insomnia walk slower, demonstrate poorer cognitive performance, and report difficulty walking or performing stairs without resting.^7,8^ Additionally, poor sleep quality has been associated with difficulties with community ambulation, navigating stairs, and performing activities of daily living.^9–12^ While there is evidence that poor sleep negatively impacts physical activity^13,14^ and physical performance,^15–17^ it remains unclear if sleep impacts mobility in particular.

Mobility is often impaired after injury or disease onset, and thus is a frequent target for rehabilitation. Understanding how sleep impacts mobility would provide insight on whether improving sleep should also be a goal during rehabilitation to enhance outcomes and recovery. Therefore, the purpose of this scoping review is to review the literature on if sleep impacts mobility in adults. We hypothesize that sleep would be positively associated with mobility, meaning better sleep would be associated with better mobility or worse sleep would be associated with worse mobility.

## METHODS

Optimal mobility has been defined as “being able to safely and reliably go where you want to go, when you want to go, and how you want to get there.”^18^ This refers to all forms of movement including basic ambulation, exercising, completion of daily responsibilities, driving, and/or using any form of public transport. The International Classification of Functioning, Disability, and Health (ICF) model includes “mobility” as a subcategory within the domain of Activities and Participation.^19^ Because the ICF uses a broad description of mobility and to keep focus for this scoping review, we focused on the “Activities” domain within the ICF. Thus, for this scoping review, mobility is defined as: “changing body position (including getting into and out of lying position, sitting position, or standing position, rolling over), walking or running (regardless of distance or surface or obstacles), and going up and down stairs.”

The Preferred Reporting Items for Systematic Reviews and Meta-Analyses extension for scoping reviews (PRISMA – ScR)^20^ guidelines and the Joanna Briggs Institute’s (JBI) updated methodology for scoping reviews^21^ were followed for this scoping review. The research question was formulated based on the Population (Adult), Concept (Sleep), and Context (Mobility) framework recommended by the JBI Manual of Evidence Synthesis. A protocol was developed and registered in Open Science Forum for conducting this scoping review.^22^

We searched in the following bibliographic databases Ovid Medline, Web of Science, Embase.com, and CINAHL. Additionally, Google Scholar and hand searching were conducted to find grey literature. The pilot search strategy of this review protocol was updated and rerun in Ovid Medline on January 3rd, 2024, when the remaining searches were completed on the same date. Controlled vocabularies such as sleep, sleep deprivation, sleep disorder, mobility, mobility limitation, actigraphy etc. along with text words on the same concepts were used for searching without applying date filters. The searches have been conducted by author PV, a biomedical librarian. A complete search history from all four databases is available in Supplementary File 1.

Criteria for study inclusion were: 1. Included human adult (>18 years old) participants, 2. study design that answered the scoping review question (i.e. cross-sectional, longitudinal, cohort, or retrospective study design; intervention if sleep was the intervention target), 3. assessed sleep as part of the study (via self-report or objective measures; assessment of “daytime sleepiness” included), 4. assessed mobility (via self-report or objective measures) per scoping review’s operational definition, 5. included statistical analyses to relate sleep assessment(s) to mobility assessment(s), and 6. included original data. Studies were excluded if: 1. they included animals, 2. they were written in a language other than English, 3. the study design did not answer the research question (i.e., reviews, opinions, editorials, protocols; studies that quantified or categorized physical activity or physical fitness without assessment of mobility per scoping review definition of mobility; studies that assessed physical activity via questionnaire such as the IPAQ; studies that assessed falls or balance; tangential measures of sleep such as heart rate variability during sleep, use of sleep medications, and risk of a sleep disorder), 4. data were reported in another study (duplicate dataset), 5. they were abstracts or non-peer reviewed sources (i.e. conference proceedings, preprint), 6. they were irrelevant to the study question.

Citations were uploaded to EndNote version 21, and duplicates were removed. Citations were then uploaded into Covidence (Veritas Health Innovation, Melbourne, Australia; www.covidence.org), a web-based collaboration software platform, to perform the review process and data extraction. Each title/abstract was independently reviewed by two of five reviewers (C.S., M.B., S.D., J.W., A.G.) and the full-text articles were reviewed by two of six reviewers (C.S., M.B., S.D., J.W., A.G., G.B.) to determine eligibility. Discrepancies were resolved by another reviewer (S.C.).

A standard data extraction form was built in Covidence by C.S. and reviewed and approved by all study personnel before data extraction started. For each article, data were extracted by two of six reviewers (C.S., M.B., S.D., J.W., A.G., G.B.) and were then reviewed for accuracy, completeness, and to reach consensus by another reviewer (S.C.). Data extracted included the first author’s last name, year the article was published, the country the study was completed, the study design, the study aim, participant information (i.e., number of participants enrolled, comorbid conditions, age, sex, race, ethnicity, comorbid conditions), information about the intervention (if applicable), the mobility outcome and classification, the sleep outcome, and main results of how sleep impacts mobility. Bias risk and quality were not assessed in accordance with updated methodological recommendations.^21^

Descriptive statistics (i.e. number and percentage) were calculated. Sleep outcomes were coded by A.G. as “self-report” (i.e. questionnaire, answer to verbal or written question, etc.) or “objective” (i.e. wearable, actigraphy, polysomnography). Mobility outcomes were coded by A.G. into seven mobility categories per our operation definition of mobility (i.e. getting into and out of lying position, getting into and out of sitting position, getting into and out of standing position, rolling over, walking, running, and going up and down stairs). The association between sleep and mobility was coded as “positive” (i.e. better sleep was associated with better mobility or poor sleep was associated with worse mobility), “negative” (i.e. better sleep was associated with worse mobility or poor sleep was associated with better mobility), “not associated” (sleep was not associated with mobility), or “mixed” (some results within the study were positive or negative and some results within the study were not associated). One author (M.B.) coded the association between sleep and mobility and the decision was reviewed by two other authors (A.G., S.C.) and consensus was reached by C.S.

## Results

The search strategy generated 8,772 references. Duplicates (n = 3,245) were removed, leaving 5,527 titles/abstracts for review. Of the titles/abstracts reviewed, 4,830 were excluded due to not meeting eligibility criteria, leaving 697 full-text articles for review. Of the full-text articles, 590 were excluded for not meeting eligibility criteria, leaving 107 eligible articles for inclusion in the scoping review (Figure 1; Supplementary File 2 Full Reference List of Included Articles).

**Figure 1.**
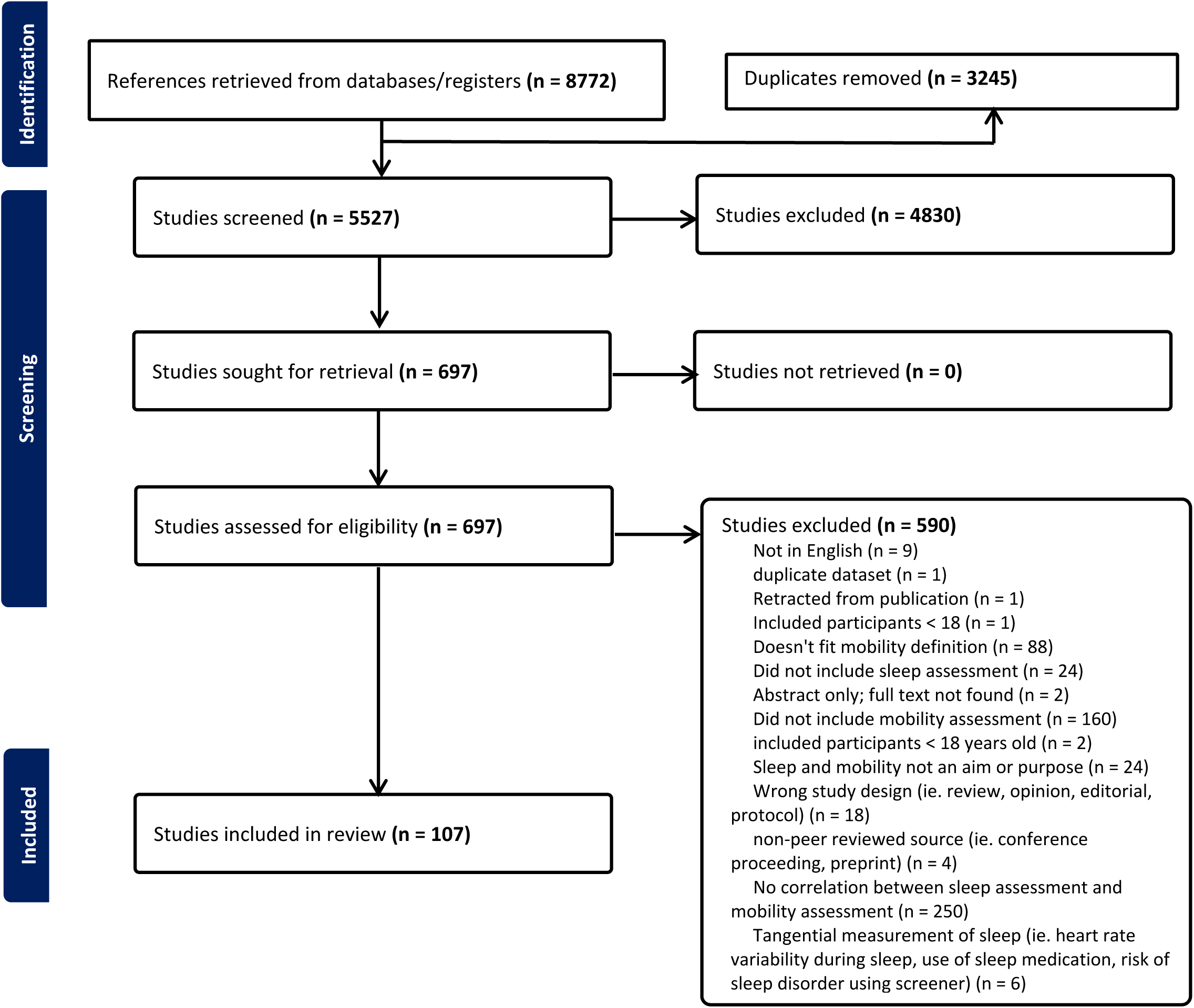

The United States was the country where the highest number of studies were conducted (n = 34, 32%) followed by China (n = 11, 10%) and Japan (n = 10, 9%; Table 1). Fifty-nine (55%) studies included people with health conditions, with neurological conditions (n = 22) being the most common category included and stroke being the most common specific neurologic condition (n = 9; Table 2). Most studies included only self-reported sleep (n = 68, 64%; Table 3) using measures ranging from 1-item questions to validated questionnaires (Supplementary Table of Data Extraction). The most common mobility category included in studies was walking (n = 101) followed by getting into and out of a sitting position (n = 26) and standing position (n = 24; Table 3). It should be noted that some studies included more than one measure to assess a mobility category (e.g. used accelerometer and a self-report question to assess walking), included more than one mobility category so included multiple mobility measures (e.g., included accelerometer to assess walking mobility and included stairs to assess stair mobility), or some measures were used to examine more than one mobility category (e.g., the 5 time sit-to-stand test was counted for both getting into and out of sitting and getting into and out of standing position).

**Table 1.** Country study was conducted organized by continent. One study included participants from two countries (United States and Iceland) so n=108.

|  | **n** |
| --- | --- |
| **Africa** | **1** |
| Nigeria | 1 |
| **Asia** | **36** |
| China | 11 |
| India | 1 |
| Japan | 10 |
| Korea | 5 |
| South Korea | 2 |
| Taiwan | 5 |
| Turkey | 2 |
| **Europe** | **23** |
| Croatia | 1 |
| Denmark | 1 |
| Finland | 1 |
| Germany | 1 |
| Greece | 2 |
| Iceland* | 1 |
| Ireland | 2 |
| Italy | 1 |
| Netherlands | 1 |
| Norway | 1 |
| Portugal | 1 |
| Spain | 4 |
| Sweden | 1 |
| United Kingdom | 5 |
| **North America** | **39** |
| Canada | 5 |
| United States* | 34 |
| **South America** | **7** |
| Brazil | 7 |
| **Australia/New Zealand** | **2** |
| Australia | 1 |
| New Zealand | 1 |

**Table 2.** Number of studies that included participants with identified health conditions organized by major category or system.

|  | n |
| --- | --- |
| **Cardiovascular Disease** | **8** |
| **Kidney Disease** | **3** |
| **Mental Health** | **2** |
| PTSD | 1 |
| Schizophrenia | 1 |
| **Metabolic** | **4** |
| Type 2 diabetes | 1 |
| Overweight or obesity | 2 |
| Prediabetes | 1 |
| **Musculoskeletal** | **3** |
| Knee Osteoarthritis | 1 |
| Fracture (trunk or lower extremity) | 1 |
| Axial Spondyloarthritis | 1 |
| **Neurological** | **22** |
| ALS | 1 |
| Cerebral Palsy | 1 |
| Parkinson's Disease | 6 |
| Stroke | 9 |
| Multiple Sclerosis | 1 |
| Acquired Brain Injury | 1 |
| Cognitive Impairment or Alzheimer's Disease | 2 |
| **Other** | **8** |
| Cancer | 4 |
| Down Syndrome | 1 |
| Pregnancy | 1 |
| Unilateral or bilateral cataract surgery | 1 |
| Physical disability | 1 |
| **Pulmonary** | **6** |
| COPD | 3 |
| Cystic fibrosis | 1 |
| Idiopathic pulmonary fibrosis | 1 |
| Interstitial lung disease | 1 |
| **Sleep Disorders** | **3** |
| Sleep Apnea | 2 |
| Insomnia | 1 |
| **Total** | **59** |

**Table 3.**
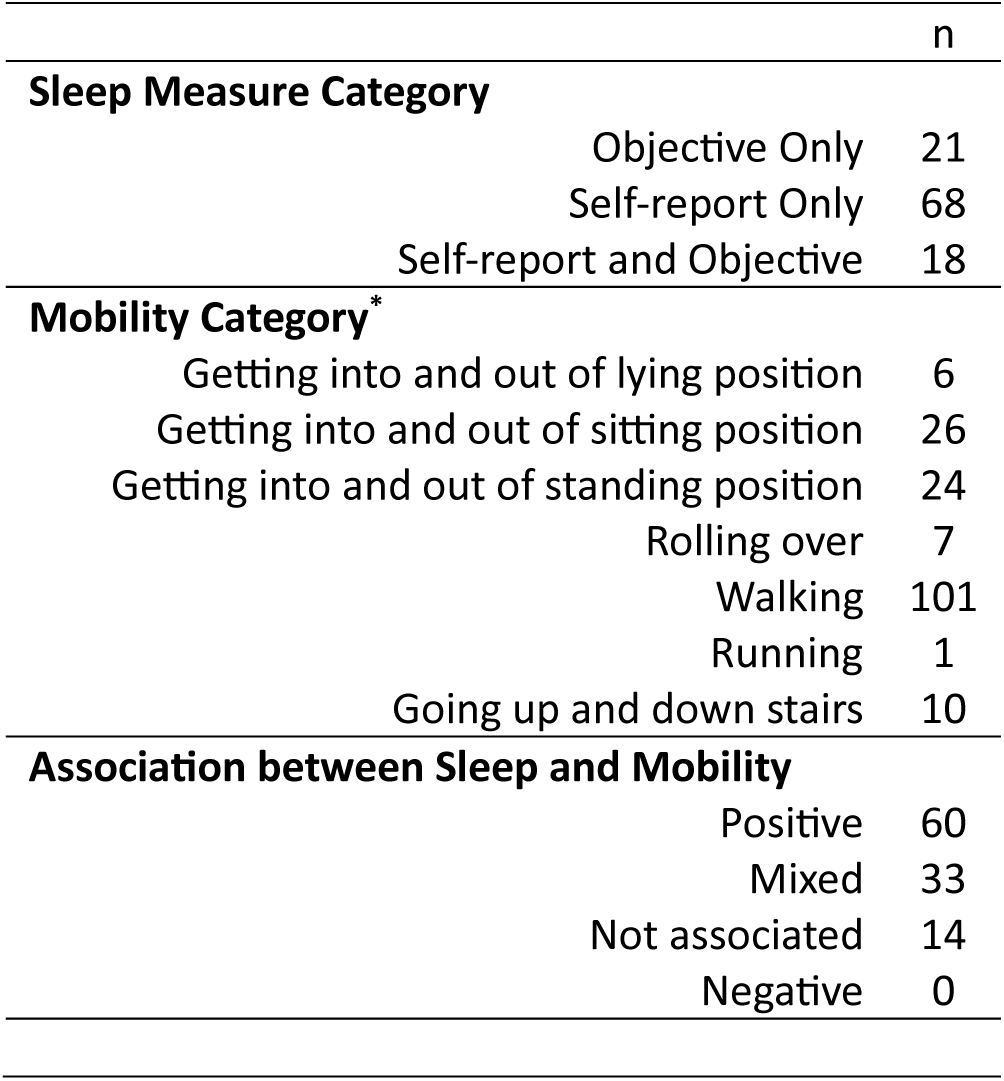
Sleep measure category, mobility category, and association between sleep and mobility. *some study included more than one mobility outcome or a mobility outcome could fall within more than one mobility category so n does not equal 107.

The majority of studies (n = 60; 56%; Table 3) reported a positive association between sleep and mobility, indicating better sleep was associated with better mobility or worse sleep was associated with worse mobility, 33 studies (31%) reported a mixed association meaning some results within the study showed a positive association while other results within the study did not show an association, and 14 studies (13%) reported no association between sleep and mobility (Supplementary Table of Data Extraction). Of the 14 studies that reported no association between sleep and mobility, n =6 did not enroll people with specific concurrent health conditions and n = 8 were spread across different health conditions (n = 2 for musculoskeletal condition, neurological condition, and “Other; n = 1 for cardiovascular and pulmonary disease; Table 4). No studies reported a negative association indicating better sleep was associated with worse mobility or worse sleep was associated with better mobility.

**Table 4.** The association between sleep and mobility by major health category or system.

|  | **n** |
| --- | --- |
| **Cardiovascular Disease** | **8** |
| Positive | 4 |
| Mixed | 3 |
| Not associated | 1 |
| **Kidney Disease** | **3** |
| Positive | 2 |
| Mixed | 1 |
| Not associated | 0 |
| **Metabolic** | **4** |
| Positive | 3 |
| Mixed | 1 |
| Not associated | 0 |
| **Mental Health** | **2** |
| Positive | 2 |
| Mixed | 0 |
| Not Associated | 0 |
| **Musculoskeletal** | **3** |
| Positive | 1 |
| Mixed | 0 |
| Not associated | 2 |
| **Neurological** | **22** |
| Positive | 14 |
| Mixed | 6 |
| Not associated | 2 |
| **Other** | **8** |
| Positive | 3 |
| Mixed | 3 |
| Not associated | 2 |
| **Pulmonary** | **6** |
| Positive | 4 |
| Mixed | 1 |
| Not associated | 1 |
| **Sleep Disorders** | **3** |
| Positive | 1 |
| Mixed | 2 |
| Not associated | 0 |

When considering the association between sleep and mobility by health condition (Table 4), most studies including people with cardiovascular, kidney, metabolic, mental health, neurological, and pulmonary conditions reported a positive association between sleep and mobility. When the number of mixed association results are combined with positive association results, between 50% (musculoskeletal conditions on the low end) to 100% (kidney, metabolic, mental health, and sleep conditions on high end) of studies reported some positive association between sleep and mobility.

When considering the association between sleep and mobility by type of sleep assessment (self-report only, objective only, both self-report and objective; Table 5), there was a higher percentage of studies that reported a positive association between sleep outcomes and mobility outcomes in studies that used only self-report sleep assessments (59%) and those that used both self-report and objective sleep assessment (56%) than studies that used objective sleep assessments only (43%). When the number of mixed association results are combined with positive association results, 81% of objective only studies, 86% of self-report only, and 100% that used both sleep assessment reported some positive association between sleep and mobility.

**Table 5.** The association between sleep and mobility by type of sleep assessment.

| Type of Sleep Assessment | n | % |
| --- | --- | --- |
| **Self-Report** | **70** |  |
| Positive | 41 | 59 |
| Mixed | 19 | 27 |
| Not associated | 10 | 14 |
| **Objective** | **21** |  |
| Positive | 9 | 43 |
| Mixed | 8 | 38 |
| Not associated | 4 | 19 |
| **Both** | **16** |  |
| Positive | 9 | 56 |
| Mixed | 7 | 44 |
| Not associated | 0 | 0 |

When considering the association between sleep and mobility by mobility category (Table 6), half or more of articles including the mobility category of getting into and out of a lying position, rolling over, walking, and going up and down stairs reported a positive association between sleep and mobility. The mobility category with the highest percentage of articles that reported no association between sleep and mobility was walking (n=14; 19%) followed by getting into and out of a standing position (n=4; 17%).

**Table 6.** The association between sleep and mobility by mobility category. Note n is greater than total number of articles included because some articles included more than one mobility category.

| **Mobility Category** | **n** | **%** |
| --- | --- | --- |
| **Getting into and out of lying position** | **6** |  |
| Positive | 3 | 50 |
| Mixed | 3 | 50 |
| Not associated | 0 | -- |
| **Getting into and out of sitting position** | **26** |  |
| Positive | 9 | 35 |
| Mixed | 13 | 50 |
| Not associated | 4 | 15 |
| **Getting into and out of standing position** | **24** |  |
| Positive | 9 | 38 |
| Mixed | 11 | 46 |
| Not associated | 4 | 17 |
| **Rolling over** | **7** |  |
| Positive | 4 | 57 |
| Mixed | 3 | 43 |
| Not associated | 0 | -- |
| **Walking** | **101** |  |
| Positive | 57 | 56 |
| Mixed | 30 | 30 |
| Not associated | 14 | 19 |
| **Running** | **1** |  |
| Positive | 0 | -- |
| Mixed | 1 | 100 |
| Not associated | 0 | -- |
| **Going up and down stairs** | **10** |  |
| Positive | 7 | 70 |
| Mixed | 2 | 20 |
| Not associated | 1 | 10 |

## Discussion

Overall, the hypothesis that sleep would be positively associated with mobility was supported. When examining the association between sleep and mobility by comorbid condition, the lowest percentage of studies that included at least some positive association between sleep and mobility (mixed + positive in Table 4) was in studies including people with musculoskeletal comorbid conditions (50% of studies) and the highest percentage (100%) was in studies including people with kidney disease, metabolic conditions, mental health conditions, and sleep disorders.

It is perhaps not a surprise that all studies including people with kidney disease, metabolic conditions, mental health conditions, and sleep disorders had at least some positive association between sleep and mobility as sleep disorders and disturbances increase risk of metabolic conditions, kidney disease, and mental health conditions. Sleep and circadian rhythms play a critical role in regulating daily physiological patterns essential for maintaining normal metabolic function. Disruption of circadian rhythms has been increasingly identified as a significant factor contributing to impaired physiological processes and the development of various diseases, particularly metabolic dysregulation.^23–25^ Factors such as irregular sleep patterns, insomnia, sleep apnea, narcolepsy, and shift work disorder are well-recognized as contributors to sleep deficiencies, which may, in turn, promote the onset of metabolic disorders.^26–29^ A growing body of evidence indicates that sleep disturbances may also contribute to the development of kidney disease. This connection is thought to arise from an inflammatory milieu and sympathetic activation within the renal vascular bed, which can damage the glomerular basement membrane and kidney tubular apparatus.^30–33^ A meta-analysis identified an association between short sleep duration and proteinuria, a surrogate marker for kidney disease progression.^34^ Additionally, renal hyperfiltration, an early indicator of renal damage, has been linked to both short sleep duration (less than 6 hours) and long sleep duration (more than 10 hours).^35,36^ Renal hyperfiltration can cause an increase in urination due to the larger volume of urine being produced by the kidneys^37^; this may impact sleep quality and an individual’s ability to get comfortable in bed and fall back asleep after getting up to use the bathroom during the night. Further, sleep disorders often co-exist with mental health conditions, and sleep disruption may be involved in the development and maintenance of mental health conditions.^38^ The mechanisms underpinning how sleep disturbances may contribute to the development of mental health disorders are likely to be a combination of circadian disruption or misalignment, genetic associations, sleep’s impact on neuroplasticity, and sleep’s impact on cognition and emotional regulation.^38^ Indeed, interventions which address sleep and circadian disruption have a positive impact on mental health conditions.^38^

It is also not a surprise that studies including people with musculoskeletal comorbid conditions had the lowest percentage of results with positive or mixed association. People with musculoskeletal comorbid conditions may have limited mobility due to the musculoskeletal condition itself (i.e., impaired joint mobility, muscle weakness, impaired mechanics, or pain). Also, only three studies included people with musculoskeletal conditions, so more research is needed in this area before conclusions can be made.

It is interesting that the majority of studies examined walking mobility (n = 101), and relatively few examined rolling over (n = 7). Humans change position, including rolling over, periodically throughout the night while sleeping, often without waking. If someone has impaired ability to roll in bed, it is conceivable that additional effort or exertion to roll or inability to roll into a more comfortable position would increase likelihood of awakening and sleep disturbances. Indeed, impaired rolling ability was associated with poorer sleep outcomes in people with Parkinson’s Disease, spinal cord injury, and amyotrophic lateral sclerosis in studies included in this scoping review.^39–42^ Similarly, getting into and out of a lying position and standing position is needed to rise from bed to go to the bathroom. More effort or exertion or frustration with impaired mobility of getting into and out of a lying or standing position could conceivably increase arousal and impair ability to return to sleep, and thus, increase sleep disturbances. Additional research is needed to examine how impaired mobility, particularly bed mobility, may impact sleep and if focused interventions to improve bed mobility also improve sleep outcomes.

A challenge with examining how sleep impacts mobility for this scoping review is the variety of measures used to assess both sleep and mobility, which made synthesis of results and comparison across studies difficult. The majority of included studies (64%) used only self-reported measures to assess sleep, and some used invalid and unreliable measures t to assess sleep or only used a single question to assess sleep. Self-report and objective sleep measures assess different sleep constructs, so it is ideal to include both types of valid and reliable measures (only 17% of included studies included both) as well as intentionally select sleep measures for the intended sleep construct (i.e., use the PSQI to assess self-reported sleep quality). Also, attempts to build consensus for mobility measures have focused on addressing the variability and lack of standardization in definitions, constructs, and assessments. Researchers emphasize the need for a unified approach to define mobility comprehensively, encompassing constructs like physical capacity, performance, and functional independence. Efforts to standardize include the development of core sets of outcome measurement instruments and detailed protocols to ensure consistency in assessment across studies. These initiatives aim to improve comparability of data, enhance the precision of findings, and facilitate large-scale data sharing, enabling a deeper understanding of complex relationships (such as those between sleep and mobility). Standardization is essential to advancing personalized and effective interventions in clinical and research contexts.^43,44^

A strength of this scoping review is that included studies were conducted in 27 different countries, spanning 6 continents (Antarctica not included), indicating generalizability of the results. However, a high number of studies (n = 40, 37%) were conducted in North America with the United States being the most represented country (n = 35, 32%). Another strength is the variety of comorbid conditions included which allowed for the exploration of how sleep impacts mobility by major health category or system. Due to the relatively small number of studies of each health category or system and the variety of specific conditions within each category/system, conclusions cannot be drawn at this point. However, this scoping review does provide insight into health categories/systems where further research is warranted. Given that most sleep disorders are treatable, addressing sleep disturbances may play a crucial role in alleviating mobility impairments in individuals with these comorbidities. Targeted interventions for sleep disorders could offer a promising pathway to improving overall mobility and quality of life in affected populations. We also acknowledge that our definition of mobility which was based on the “mobility” subcategory within the domain of Activities and Participation of the ICF model^19^ excludes other types of mobility such as wheelchair mobility, driving, jumping, climbing, swimming, and use of transportation. Future reviews should consider examining how sleep impacts mobility of other classification of mobility.

Overall, this scoping review provides a description of the literature on the association between sleep and mobility in adults. The results indicate that there is a positive association between sleep and mobility; better sleep is associated with better mobility and worse sleep is associated with worse mobility, particularly for getting into and out of lying position, rolling over, walking, and going up and down stairs. Due to the variety of sleep and mobility outcomes used, it was challenging to compare studies and synthesize results. Most studies including people with cardiovascular disease, kidney disease, metabolic conditions, mental health conditions, neurological conditions, and pulmonary disease reported a positive association between sleep and mobility. However, due to a relatively small sample size within a given health condition, conclusions cannot be drawn, and further research is needed.

## Data Availability

All data produced in the present study are available upon reasonable request to the authors

## Conflict of interest

Catherine Siengsukon: Owner and CEO of Sleep Health Education, LLC

Mahya Beheshti: None to declare

Sarah J. Donkers: None to declare

Silvana L. Costa: None to declare

Prasanna Vaduvathiriyan: None to declare

Allison Glaser: None to declare

Garrett Baber: None to declare

Joy Williams: None to declare

## Notes

### Funding Statement

This study did not receive any funding

