## Supplementary File for "Does sleep impact mobility in adults? A scoping review"

Ovid MEDLINE(R) and Epub Ahead of Print, In-Process, In-Data-Review & Other Non-Indexed Citations, Daily and Versions <1946 to January 02, 2024>

| # | Query | Results |
|---|---|---|
| 1 | (sleep disorder$ or sleep defici$ or sleep qualit$ or sleep behavior$ or sleep dysfunction$ or insomnia$ or sleepless$ or dyssomnia or parasomnia$ or somnolence).ti,ab,kw. | 84,233 |
| 2 | (mobility or mobilize or mobilise or mobilization or mobilisation).ti,ab,kw. | 250,945 |
| 3 | 1 and 2 | 437 |
| 4 | ((functional mobilit$ or functional abilit$ or mobility or physical performanc$ or physical function$ or physical activit$) adj3 sleep$).ti. | 763 |
| 5 | exp Sleep Wake Disorders/ or sleep deprivation/ or sleep duration/ or sleep quality/ or sleep latency/ or sleep stages/ or sleep/ | 166,839 |
| 6 | 1 or 5 | 201,872 |
| 7 | mobility limitation/ | 5,299 |
| 8 | 6 and 7 | 59 |
| 9 | walking speed/ or gait analysis/ or gait/ or locomotion/ or "activities of daily living"/ or functional status/ or accelerometry/ or actigraphy/ | 148,738 |
| 10 | (co or pp or ep).fs. | 5,596,309 |
| 11 | 9 and 10 | 52,171 |
| 12 | 6 and 11 | 2,439 |
| 13 | ((walking speed$ or gait speed$) and sleep$).ti,ab. | 154 |
| 14 | 3 or 4 or 8 or 12 or 13 | 3,689 |
| 15 | 14 not ((exp infant/ or exp child/ or adolescent/) not exp adult/) | 3,188 |
| 16 | 15 not (exp animals/ not humans.sh.) | 3,096 |

**Embase**

| | | |
|---|---|---|
| #16 | #15 AND 'Article'/it | 2407 |
| #15 | #14 NOT (('adolescent'/exp OR 'child'/exp OR child*:ti,ab,kw OR youth:ti,ab,kw OR teen:ti,ab,kw) NOT ('adult'/exp OR 'aged'/exp OR 'middle aged'/exp OR adult*:ti,ab,kw)) | 4253 |
| #14 | #13 NOT ([animals]/lim NOT [humans]/lim) | 4949 |
| #13 | #3 OR #4 OR #10 OR #12 | 5042 |
| #12 | #10 OR #11 | 2645 |
| #11 | ('walking speed':ti,ab,kw OR 'gait speed':ti,ab,kw) AND sleep:ti,ab,kw | 256 |
| #10 | #8 AND #9 | 2418 |
| #9 | 'walking'/exp/mj OR 'gait disorder'/mj OR 'immobility'/mj OR 'limited mobility'/mj OR 'walking difficulty'/mj OR 'agility'/mj OR 'physical mobility'/mj OR 'movement (physiology)'/mj OR 'daily life activity'/mj OR 'functional status'/mj OR 'functional status assessment'/exp/mj OR 'actimetry'/mj | 106247 |

| | | |
|---|---|---|
| #8 | 'sleep disorder'/exp/mj OR 'sleep deprivation'/exp/mj OR 'sleep parameters'/exp/mj OR 'sleep'/exp/mj OR #1 | 282634 |
| #7 | #5 AND #6 | 39548 |
| #6 | 'walking'/exp OR 'gait disorder'/de OR 'limited mobility'/de OR 'walking difficulty'/de OR 'agility'/de OR 'physical mobility'/de OR 'movement (physiology)'/de OR 'daily life activity'/de OR 'functional status'/de OR 'functional status assessment'/exp OR 'actimetry'/de | 627895 |
| #5 | 'sleep disorder'/exp OR 'sleep deprivation'/exp OR 'sleep parameters'/exp OR 'sleep'/exp OR #1 | 514144 |
| #4 | ('functional mobilit*':ti OR 'functional abilit*':ti OR 'physical performanc*':ti OR 'mobility limitation*':ti OR 'physical activit*':ti) AND sleep*:ti | 1728 |
| #3 | #1 AND #2 | 787 |
| #2 | mobility:ti,ab,kw OR mobilize:ti,ab,kw OR mobilise:ti,ab,kw OR mobilization:ti,ab,kw OR mobilisation:ti,ab,kw | 309634 |
| #1 | 'sleep disorder*':ti,ab,kw OR 'sleep defici*':ti,ab,kw OR 'sleep qualit*':ti,ab,kw OR 'sleep behavior*':ti,ab,kw OR 'sleep dysfunction*':ti,ab,kw OR insomnia*:ti,ab,kw OR sleepless*:ti,ab,kw OR parasomnia*:ti,ab,kw OR somnolence:ti,ab,kw | 137260 |

CINAHL

| | | | |
|---|---|---|---|
| S14 | S12 NOT ( ( (MH "Animals") OR (MH "Animal Studies") ) NOT (MH "Human") ) | Limiters - English Language; Publication Type: Journal Article Expanders - Apply equivalent subjects Search modes - Boolean/Phrase | 3,097 |
| S13 | S12 NOT ( ( (MH "Animals") OR (MH "Animal Studies") ) NOT (MH "Human") ) | Expanders - Apply equivalent subjects Search modes - Boolean/Phrase | 3,262 |
| S12 | S11 NOT ( ( MH "Child+" OR MH "Adolescence" ) NOT MH "Adult+" )) | Expanders - Apply equivalent subjects Search modes - Boolean/Phrase | 3,295 |
| S11 | S3 OR S4 OR S7 OR S9 OR S10 | Expanders - Apply equivalent subjects Search modes - Boolean/Phrase | 3,860 |
| S10 | ( TI ( walking speed or gait speed ) OR AB ( walking speed or gait speed ) ) AND ( TI sleep* OR AB sleep ) | Expanders - Apply equivalent subjects Search modes - Boolean/Phrase | 66 |
| S9 | S5 AND S8 | Expanders - Apply equivalent subjects Search modes - Boolean/Phrase | 2,637 |
| S8 | ( ( ( (MH "Activities of Daily Living") OR (MH "Functional Status") ) OR ( (MH "Mobility (Iowa NOC)") OR (MH "Mobility | Expanders - Apply equivalent subjects | 73,943 |

| | | | |
|---|---|---|---|
| | Level (Iowa NOC)") OR (MH "Impaired Physical Mobility (NANDA)") OR (MH "Physical Mobility Impairment (Saba CCC)") ) ) OR ( (MH "Accelerometry") OR (MH "Actigraphy") ) ) OR TI ( "walking speed" or "gait speed" or "gait analysis" ) OR AB ( walking speed" or "gait speed" or "gait analysis" ) | Search modes - Boolean/Phrase | |
| S7 | S5 AND S6 | Expanders - Apply equivalent subjects Search modes - Boolean/Phrase | 506 |
| S6 | TI "mobility limitation" OR AB "mobility limitation" OR (MM "Movement+") | Expanders - Apply equivalent subjects Search modes - Boolean/Phrase | 63,059 |
| S5 | ( ( (MH "Sleep") OR (MH "Sleep Deprivation") OR (MH "Sleep Duration") OR (MH "Sleep Quality") OR (MH "Sleep Latency") OR (MH "Deep Sleep") OR (MH "Sleep Stages") OR (MH "Sleep, REM") ) OR ( (MH "Sleep Disorders") OR (MH "Sleep Deprivation") ) ) OR ( TI ( sleep disorder* OR sleep defici* OR sleep quality* or sleep behavior* OR "sleep dysfunction*" OR insomnia* OR sleepless* OR dyssomnia OR parasomnia* OR somnolence ) OR AB ( "sleep disorder*" OR "sleep defici*" OR "sleep quality*" or "sleep behavior*" OR "sleep dysfunction*" OR insomnia* OR sleepless* OR dyssomnia OR parasomnia* OR somnolence ) | Expanders - Apply equivalent subjects Search modes - Boolean/Phrase | 61,128 |
| S4 | ( TI ( "functional mobilit*" OR "functional ability*" OR mobility OR "physical performance*" ) OR AB ( "functional mobilit*" OR "functional ability*" OR mobility OR "physical performance*" ) ) AND ( ( TI sleep* OR AB sleep* ) ) | Expanders - Apply equivalent subjects Search modes - Boolean/Phrase | 832 |
| S3 | S1 AND S2 | Expanders - Apply equivalent subjects Search modes - Boolean/Phrase | 175 |
| S2 | TI ( mobility or mobilize or mobilise or mobilization or mobilisation ) OR AB ( mobility or mobilize or mobilise or mobilization or mobilisation ) | Expanders - Apply equivalent subjects Search modes - Boolean/Phrase | 48,175 |
| S1 | TI ( sleep disorder* OR sleep defici* OR sleep quality* or sleep behavior* OR sleep dysfunction* OR insomnia* OR sleepless* OR dyssomnia OR parasomnia* OR somnolence ) OR AB ( sleep disorder* OR sleep defici* OR sleep quality* or sleep behavior* OR sleep dysfunction* OR insomnia* OR sleepless* OR dyssomnia OR parasomnia* OR somnolence ) | Expanders - Apply equivalent subjects Search modes - Boolean/Phrase | 31,190 |

### Full Reference List of Included Articles


1. Ahern, T., O'Malley, E., Dunlevy, C., Khattak, A., O'Brien, H., Hassan, T., Cusack, T., McNicholas, W. T., & O'Shea, D. (2020). Sleep duration and physical function in people with severe obesity: a prospective cross-sectional study. *Irish Journal of Medical Science*, *189*(2), 517-523.
2. Auyeung, T. W., Kwok, T., Leung, J., Lee, J. S., Ohlsson, C., Vandenput, L., Wing, Y. K., & Woo, J. (2015). Sleep Duration and Disturbances Were Associated With Testosterone Level, Muscle Mass, and Muscle Strength--A Cross-Sectional Study in 1274 Older Men. *Journal of the American Medical Directors Association*, *16*(7), 630.e631-636.
3. Awotidebe, T. O., Adeyeye, V. O., Adedoyin, R. A., Ogunyemi, S. A., Oke, K. I., Ativie, R. N., Adeola, G. B., Akindele, M. O., & Balogun, M. O. (2017). Assessment of functional capacity and sleep quality of patients with chronic heart failure. *Hong Kong Physiotherapy Journal*, *36*, 17-24.
4. Ayaki, M., Negishi, K., & Tsubota, K. (2014). Rejuvenation effects of cataract surgery with ultraviolet blocking intra-ocular lens on circadian rhythm and gait speed. *Rejuvenation Research*, *17*(4), 359-365.
5. Batalla-Martin, D., Belzunegui-Eraso, A., Miralles Garijo, E., Martinez Martin, E., Romani Garcia, R., Heras, J. S. M., Lopez-Ruiz, M., & Martorell-Poveda, M. A. (2020). Insomnia in Schizophrenia Patients: Prevalence and Quality of Life. *International Journal of Environmental Research & Public Health [Electronic Resource]*, *17*(4), 19.
6. Benito-Villena, R., Guerrero-Martinez, I., Naveiro-Fuentes, M., Cano-Ibanez, N., Femia-Marzo, P., Gallo-Vallejo, J. L., Mozas-Moreno, J., & Amezcua-Prieto, C. (2022). Walking Promotion in Pregnancy and Its Effects on Insomnia: Results of Walking_Preg Project (WPP) Clinical Trial. *International Journal of Environmental Research & Public Health [Electronic Resource]*, *19*(16), 13.
7. Bernstein, John P. K., De Vito, A., Weitzner, Daniel S., MacAulay, R., Calamia, M., Brouillette, R., Foil, H., Carmichael, Owen T., Keller, Jeffrey N., Bernstein, J. P. K., De Vito, A., Weitzner, D. S., Carmichael, O. T., & Keller, J. N. (2020). Examining Relationships between Multiple Self-Reported Sleep Measures and Gait Domains in Cognitively Healthy Older Adults. *Gerontology*, *66*(1), 47-54.
8. Bertapelli, F., Johnson, M., Pitetti, K., Smith, M. C., Carlson, B., Curtis, J. S., & Agiovlasitis, S. (2022). Association between sleep quality and physical functioning in adults with Down syndrome: A brief report. *Disability & Health Journal*, *15*(1), 101173.
9. Boolani, A., Martin, J., Huang, H., Yu, L. F., Stark, M., Grin, Z., Roy, M., Yager, C., Teymouri, S., Bradley, D., Martin, R., Fulk, G., & Kakar, R. S. (2022). Association between Self-Reported Prior Night's Sleep and Single-Task Gait in Healthy, Young Adults: A Study Using Machine Learning. *Sensors*, *22*(19), 29.
10. Chasens, E. R., & Yang, K. (2012). Insomnia and Physical Activity in Adults With Prediabetes. *Clinical Nursing Research*, *21*(3), 294-308.
11. Chen, H., Zhang, G., Wang, Z., Feng, S., & Li, H. (2022). The Associations between Daytime Physical Activity, While-in-Bed Smartphone Use, Sleep Delay, and Sleep Quality: A 24-h Investigation among Chinese College Students. *International Journal of Environmental Research and Public Health*, *19*(15).

12. Chen, L. J., Fox, K. R., Sun, W. J., Tsai, P. S., Ku, P. W., & Chu, D. (2018). Associations between walking parameters and subsequent sleep difficulty in older adults: A 2-year follow-up study. *Journal of Sport & Health Science*, *7*(1), 95-101.
13. Cheng, H.-P., Chen, C.-H., Lin, H.-S., Wang, J.-J., & Yen, M. (2022). Relationship between Walking Activity and Sleep Quality among Community-Dwelling Older Adults. *Journal of Community Health Nursing*, *39*(2), 127-138.
14. Chevance, G., Baretta, D., Romain, A. J., Godino, J. G., & Bernard, P. (2022). Day-to-day associations between sleep and physical activity: a set of person-specific analyses in adults with overweight and obesity. *Journal of Behavioral Medicine*, *45*(1), 14-27.
15. Chien, M. Y., & Chen, H. C. (2015). Poor sleep quality is independently associated with physical disability in older adults. *Journal of Clinical Sleep Medicine*, *11*(3), 225-232.
16. Cole, C. S., Richards, K. C., Beck, C. C., Roberson, P. K., Lambert, C., Furnish, A., Free, J., & Tackett, J. (2009). Relationships among disordered sleep and cognitive and functional status in nursing home residents. *Research in Gerontological Nursing*, *2*(3), 183-191.
17. Curran, M., Tierney, A. C., Button, B., Collins, L., Kennedy, L., McDonnell, C., Sheikhi, A., Jurascheck, A., Casserly, B., & Cahalan, R. (2022). Physical Activity and Sedentary Behavior in Adults With Cystic Fibrosis: Association With Aerobic Capacity, Lung Function, Sleep, Well-Being, and Quality of Life. *Respiratory Care*, *67*(3), 339-346.
18. Dam, T. T., Ewing, S., Ancoli-Israel, S., Ensrud, K., Redline, S., & Stone, K. (2008). Association between sleep and physical function in older men: the osteoporotic fractures in men sleep study. *Journal of the American Geriatrics Society*, *56*(9), 1665-1673.
19. Davis, J. C., Falck, R. S., Best, J. R., Chan, P., Doherty, S., & Liu-Ambrose, T. (2019). Examining the Inter-relations of Depression, Physical Function, and Cognition with Subjective Sleep Parameters among Stroke Survivors: A Cross-sectional Analysis. *Journal of Stroke & Cerebrovascular Diseases*, *28*(8), 2115-2123.
20. Denison, H. J., Jameson, K. A., Sayer, A. A., Patel, H. P., Edwards, M. H., Arora, T., Dennison, E. M., Cooper, C., & Baird, J. (2021). Poor sleep quality and physical performance in older adults. *Sleep Health*, *7*(2), 205-211.
21. Diaz-Abad, M., Buczyner, J. R., Venza, B. R., Scharf, S. M., Kwan, J. Y., Lubinski, B., & Russell, J. W. (2018). Poor sleep quality in patients with amyotrophic lateral sclerosis at the time of diagnosis. *Journal of Clinical Neuromuscular Disease*, *20*(2), 60-68.
22. Dobrosielski, D. A., Phan, P., Miller, P., Bohlen, J., Douglas-Burton, T., & Knuth, N. D. (2016). Associations between vasodilatory capacity, physical activity and sleep among younger and older adults. *European Journal of Applied Physiology*, *116*(3), 495-502.
23. Edgar, D. T., Gill, N. D., Beaven, C. M., Zaslona, J. L., & Driller, M. W. (2021). Sleep duration and physical performance during a 6-week military training course. *Journal of Sleep Research*, *30*(6), e13393.
24. Endeshaw, Y. W., Unruh, M. L., Kutner, M., Newman, A. B., & Bliwise, D. L. (2009). Sleep-disordered breathing and frailty in the Cardiovascular Health Study Cohort. *American Journal of Epidemiology*, *170*(2), 193-202.
25. Ezeugwu, V. E., & Manns, P. J. (2017). Sleep Duration, Sedentary Behavior, Physical Activity, and Quality of Life after Inpatient Stroke Rehabilitation. *Journal of Stroke & Cerebrovascular Diseases*, *26*(9), 2004-2012.
26. Feng, Y., Maislin, D., Keenan, B. T., Gislason, T., Arnardottir, E. S., Benediktsdottir, B., Chirinos, J. A., Townsend, R. R., Staley, B., Pack, F. M., Sifferman, A., Pack, A. I., &

Kuna, S. T. (2018). Physical Activity Following Positive Airway Pressure Treatment in Adults With and Without Obesity and With Moderate-Severe Obstructive Sleep Apnea. *Journal of Clinical Sleep Medicine*, *14*(10), 1705-1715.

27. Fex, A., Barbat-Artigas, S., Dupontgand, S., Filion, M.-E., Karelis, A. D., & Aubertin-Leheudre, M. (2012). Relationship between long sleep duration and functional capacities in postmenopausal women. *Journal of Clinical Sleep Medicine*, *8*(3), 309-313.
28. Fleming, M. K., Smejka, T., Henderson Slater, D., van Gils, V., Garratt, E., Yilmaz Kara, E., & Johansen-Berg, H. (2020). Sleep Disruption After Brain Injury Is Associated With Worse Motor Outcomes and Slower Functional Recovery. *Neurorehabilitation & Neural Repair*, *34*(7), 661-671.
29. Fongen, C., Dagfinrud, H., Bilberg, A., & Sveaas, S. H. (2023). Reduced sleep quality is highly prevalent and associated with physical function and cardiorespiratory fitness in patients with axial spondyloarthritis: a cross-sectional study. *Scandinavian Journal of Rheumatology*, 1-10.
30. Fritschi, C., Bronas, U. G., Park, C. G., Collins, E. G., & Quinn, L. (2017). Early declines in physical function among aging adults with type 2 diabetes. *Journal of Diabetes & its Complications*, *31*(2), 347-352.
31. Fulk, G., Duncan, P., & Klingman, K. J. (2020). Sleep problems worsen health-related quality of life and participation during the first 12 months of stroke rehabilitation. *Clinical Rehabilitation*, *34*(11), 1400-1408.
32. Goldman, S. E., Stone, K. L., Ancoli-Israel, S., Blackwell, T., Ewing, S. K., Boudreau, R., Cauley, J. A., Hall, M., Matthews, K. A., & Newman, A. B. (2007). Poor sleep is associated with poorer physical performance and greater functional limitations in older women. *Sleep*, *30*(10), 1317-1324.
33. Gonzalez-Sanchez, J., Recio-Rodriguez, J. I., Gomez-Marcos, M. A., Patino-Alonso, M. C., Agudo-Conde, C., & Garcia-Ortiz, L. (2019). Relationship between the presence of insomnia and walking physical activity and diet quality: A cross-sectional study in a sample of Spanish adults. *Medicina Clinica*, *152*(9), 339-345.
34. Greysen, S. R., Waddell, K. J., & Patel, M. S. (2022). Exploring Wearables to Focus on the "Sweet Spot" of Physical Activity and Sleep After Hospitalization: Secondary Analysis. *JMIR MHealth and UHealth*, *10*(4), e30089.
35. Gururaj, R., Samuel, S. R., Vijaya Kumar, K., Hegde, A., Prakash Saxena, P. U., Nagaraja, R., & Palesh, O. (2021). Relationship between physical activity, objective sleep parameters, and circadian rhythm in patients with head and neck cancer receiving chemoradiotherapy: A longitudinal study. *Laryngoscope Investigative Otolaryngology*, *6*(6), 1455-1460.
36. Hellstrom, P., Israelsson, J., Hellstrom, A., Hjelm, C., Brostrom, A., & Arestedt, K. (2023). Is insomnia associated with self-reported health and life satisfaction in cardiac arrest survivors? A cross-sectional survey. *Resuscitation Plus*, *15*, 100455.
37. Hirata, R. P., Dala Pola, D. C., Schneider, L. P., Bertoche, M. P., Furlanetto, K. C., Hernandes, N. A., Mesas, A. E., & Pitta, F. (2020). Tossing and turning: Association of sleep quantity–quality with physical activity in copd. *Erj Open Research*, *6*(4).
38. Holingue, C., Owusu, J. T., Tzuang, M., Nyhuis, C. C., Yaffe, K., Stone, K. L., Rebok, G. W., Ancoli-Israel, S., & Spira, A. P. (2023). Accelerometer-assessed sleep and decline in physical function in older men. *Sleep Health*, *23*, 23.

39. Howell, D. R., Berkstresser, B., Wang, F., Buckley, T. A., Mannix, R., Stillman, A., & Meehan, W. P., 3rd. (2018). Self-reported sleep duration affects tandem gait, but not steady-state gait outcomes among healthy collegiate athletes. *Gait & Posture*, *62*, 291-296.
40. Huang, W. C., Lin, C. Y., Togo, F., Lai, T. F., Liao, Y., Park, J. H., Hsueh, M. C., & Park, H. (2021). Association between objectively measured sleep duration and physical function in community-dwelling older adults. *Journal of Clinical Sleep Medicine*, *17*(3), 515-520.
41. Hur, S. A., Guler, S. A., Khalil, N., Camp, P. G., Guenette, J. A., & Ryerson, C. J. (2022). Characterization and determinants of sleep measured by self-report and wrist actigraphy in patients with interstitial lung disease. *Canadian Journal of Respiratory, Critical Care, and Sleep Medicine*, *6*(2), 88-96.
42. Izawa, K. P., Watanabe, S., Oka, K., Hiraki, K., Morio, Y., Kasa-hara, Y., Takeichi, N., Tsukamoto, T., Osada, N., Omiya, K., & Makuuchi, H. (2011). Relation between sleep quality and physical activity in chronic heart failure patients. *Recent Patents on Cardiovascular Drug Discovery*, *6*(3), 161-167.
43. Jean, R. E., Duttuluri, M., Gibson, C. D., Mir, S., Fuhrmann, K., Eden, E., & Supariwala, A. (2017). Improvement in Physical Activity in Persons With Obstructive Sleep Apnea Treated With Continuous Positive Airway Pressure. *Journal of Physical Activity & Health*, *14*(3), 176-182.
44. Jeon, S., & Redeker, N. S. (2016). Sleep Disturbance, Daytime Symptoms, and Functional Performance in Patients With Stable Heart Failure: A Mediation Analysis. *Nursing Research*, *65*(4), 259-267.
45. Kaizu, Y., Kasuga, T., Takahashi, Y., Otani, T., & Miyata, K. (2022). Sleep Should Be Focused on When Analyzing Physical Activity in Hospitalized Older Adults after Trunk and Lower Extremity Fractures—A Pilot Study. *Healthcare (2227-9032)*, *10*(8), 1429-N.PAG.
46. Kasovic, M., Stefan, A., & Stefan, L. (2021). The Associations Between Objectively Measured Gait Speed and Subjective Sleep Quality in First-Year University Students, According to Gender. *Nature & Science of Sleep*, *13*, 1663-1668.
47. Kataoka, H., Saeki, K., Yamagami, Y., Sugie, K., & Obayashi, K. (2020). Quantitative associations between objective sleep measures and early-morning mobility in Parkinson's disease: cross-sectional analysis of the PHASE study. *Sleep*, *43*(1), 13.
48. Kaya, T., Nalbant, A., Yildirim, I., Issever, K., Karacaer, C., Bilgin, C., Vatan, M. B., & Acar, T. (2023). Relationship between Sleep Quality and Gait Speed in Geriatric Patients. *Journal of the Liaquat University of Medical and Health Sciences*, *22*(1), 9-13.
49. Kim, M., Yoshida, H., Sasai, H., Kojima, N., & Kim, H. (2015). Association between objectively measured sleep quality and physical function among community-dwelling oldest old Japanese: A cross-sectional study. *Geriatrics & gerontology international*, *15*(8), 1040-1048.
50. Kim, Y., & Moon, H. M. (2019). Association between quality of life and sleep time among community-dwelling stroke survivors: Findings from a nationally representative survey. *Geriatrics & gerontology international*, *19*(12), 1226-1230.
51. Kline, C. E., Colvin, A. B., Pettee Gabriel, K., Karvonen-Gutierrez, C. A., Cauley, J. A., Hall, M. H., Matthews, K. A., Ruppert, K. M., Neal-Perry, G. S., Strotmeyer, E. S., & Sternfeld, B. (2021). Associations between longitudinal trajectories of insomnia

symptoms and sleep duration with objective physical function in postmenopausal women: the Study of Women's Health Across the Nation. *Sleep*, *44*(8), 13.
52. Kubala, A. G., Buysse, D. J., Brindle, R. C., Krafty, R. T., Thayer, J. F., Hall, M. H., & Kline, C. E. (2020). The association between physical activity and a composite measure of sleep health. *Sleep and Breathing*, *24*(3), 1207-1214.
53. Kurose, S., Miyauchi, T., Yamashita, R., Tamaki, S., Imai, M., Nakashima, Y., Umeda, Y., Sato, S., Kimura, Y., & Masuda, I. (2019). Association of locomotive activity with sleep latency and cognitive function of elderly patients with cardiovascular disease in the maintenance phase of cardiac rehabilitation. *Journal of Cardiology*, *73*(6), 530-535.
54. Lavoie-Tremblay, M., Sounan, C., Trudel, J. G., Lavigne, G. L., Martin, K., & Lowensteyn, I. (2014). Impact of a pedometer program on nurses working in a health-promoting hospital. *Health Care Manager*, *33*(2), 172-180.
55. Lee, P. F., Ho, C. C., Yeh, D. P., Hung, C. T., Chang, Y. C., Liu, C. C., Tseng, C. Y., & Hsieh, X. Y. (2020). Cross-Sectional Associations of Physical Fitness Performance Level and Sleep Duration among Older Adults: Results from the National Physical Fitness Survey in Taiwan. *International Journal of Environmental Research and Public Health*, *17*(2).
56. Lee, S., & Shin, C. (2017). Interaction of obstructive sleep apnoea and cognitive impairment with slow gait speed in middle-aged and older adults. *Age & Ageing*, *46*(4), 653-659.
57. Lee, T., Lee, M., Youm, C., Noh, B., & Park, H. (2020). Association between Gait Variability and Gait-Ability Decline in Elderly Women with Subthreshold Insomnia Stage. *International Journal of Environmental Research & Public Health [Electronic Resource]*, *17*(14), 17.
58. Lee, W., Jeong, S., Lee, B. S., Lim, J. C., & Kim, O. (2023). Association between functional outcomes and psychological variables in persons with spinal cord injury. *Scientific Reports*, *13*(1), 23092.
59. Lorenz, R. A., Budhathoki, C. B., Kalra, G. K., & Richards, K. C. (2014). The relationship between sleep and physical function in community-dwelling adults: a pilot study. *Family & Community Health*, *37*(4), 298-306.
60. Louter, M., Sloun, R. J. G., Pevernagie, D. A. A., Arends, J., Cluitmans, P. J., Bloem, B. R., & Overeem, S. (2013). Subjectively impaired bed mobility in Parkinson disease affects sleep efficiency. *Sleep Medicine*, *14*(7), 668-674.
61. Lü, J., Huang, L., Wu, X., Fu, W., & Liu, Y. (2017). Effect of Tai Ji Quan training on self-reported sleep quality in elderly Chinese women with knee osteoarthritis: a randomized controlled trail. *Sleep Medicine*, *33*, 70-75.
62. MacCarthy, M., Heyn, P., Tagawa, A., & Carollo, J. (2022). Walking speed and patient-reported outcomes in young adults with cerebral palsy. *Developmental Medicine & Child Neurology*, *64*(10), 1281-1288.
63. Malinowska, K. B., Okura, M., Ogita, M., Yamamoto, M., Nakai, T., Numata, T., Tsuboyama, T., & Arai, H. (2016). Effect of self-reported quality of sleep on mobility in older adults. *Geriatrics & gerontology international*, *16*(2), 266-271.
64. Mantovani, A. M., Duncan, S., Codogno, J. S., Lima, M. C., & Fernandes, R. A. (2016). Different Amounts of Physical Activity Measured by Pedometer and the Associations With Health Outcomes in Adults. *Journal of Physical Activity & Health*, *13*(11), 1183-1191.

65. Mei, Y. X., Zhang, Z. X., Wu, H., Hou, J., Liu, X. T., Sang, S. X., Mao, Z. X., Zhang, W. H., Yang, D. B., & Wang, C. J. (2022). Health-Related Quality of Life and Its Related Factors in Survivors of Stroke in Rural China: A Large-Scale Cross-Sectional Study. *Frontiers in Public Health*, *10*, 810185.
66. Moon, H. I., Yoon, S. Y., Jeong, Y. J., & Cho, T. H. (2018). Sleep disturbances negatively affect balance and gait function in post-stroke patients. *Neurorehabilitation*, *43*(2), 211-218.
67. Motohashi, Y., Maeda, A., Nakamura, K., Higuchi, S., Liu, Y., & Yuasa, T. (1999). Sleep-wake rhythm and physical fitness in relation to activities of daily living in stroke survivors residing at home. *Environmental Health and Preventive Medicine*, *3*(4), 218-222.
68. Nakakubo S, Doi T, Shimada H, Ono R, Makizako H, Tsutsumimoto K, Hotta R, Suzuki T. (2018). The Association Between Excessive Daytime Sleepiness and Gait Parameters in Community-Dwelling Older Adults: Cross-Sectional Findings From the Obu Study of Health Promotion for the Elderly. Journal of Aging & Health, 30(2), 213-228.
69. Neale, C. D., Christensen, P. E., Dall, C., Ulrik, C. S., Godtfredsen, N., & Hansen, H. (2022). Sleep Quality and Self-Reported Symptoms of Anxiety and Depression Are Associated with Physical Activity in Patients with Severe COPD. *International Journal of Environmental Research and Public Health*, *19*(24).
70. O'Dowd, S., Galna, B., Morris, R., Lawson, R. A., McDonald, C., Yarnall, A. J., Burn, D. J., Rochester, L., & Anderson, K. N. (2017). Poor Sleep Quality and Progression of Gait Impairment in an Incident Parkinson's Disease Cohort. *Journal of Parkinson's Disease*, *7*(3), 465-470.
71. Oliveira de Almeida, F., Ugrinowitsch, C., Brito, L. C., Milliato, A., Marquesini, R., Moreira-Neto, A., Barbosa, E. R., Horak, F. B., Mancini, M., & Silva-Batista, C. (2021). Poor sleep quality is associated with cognitive, mobility, and anxiety disability that underlie freezing of gait in Parkinson's disease. *Gait Posture*, *85*, 157-163.
72. Overcash, J., Tan, A., Patel, K., & Noonan, A. M. (2018). Factors Associated With Poor Sleep in Older Women Diagnosed With Breast Cancer. *Oncology Nursing Forum*, *45*(3), 359-371.
73. Pan, C. W., Cong, X., Zhou, H. J., Li, J., Sun, H. P., Xu, Y., & Wang, P. (2017). Self-Reported Sleep Quality, Duration, and Health-Related Quality of Life in Older Chinese: Evidence From a Rural Town in Suzhou, China. *Journal of Clinical Sleep Medicine*, *13*(8), 967-974.
74. Papazisis, Z., Nikolaidis, P. T., & Trakada, G. (2021). Sleep, Physical Activity, and Diet of Adults during the Second Lockdown of the COVID-19 Pandemic in Greece. *International Journal of Environmental Research & Public Health [Electronic Resource]*, *18*(14), 08.
75. Park, M., Buchman, A. S., Lim, A. S., Leurgans, S. E., & Bennett, D. A. (2014). Sleep complaints and incident disability in a community-based cohort study of older persons. *American Journal of Geriatric Psychiatry*, *22*(7), 718-726.
76. Redeker, N. S., Jeon, S., Muench, U., Campbell, D., Walsleben, J., & Rapoport, D. M. (2010). Insomnia symptoms and daytime function in stable heart failure. *Sleep*, *33*(9), 1210-1216.

77. Reimers, A. K., Heidenreich, V., Bittermann, H. J., Knapp, G., & Reimers, C. D. (2021). Accelerometer-measured physical activity and its impact on sleep quality in patients suffering from restless legs syndrome. *BMC Neurology*, *21*(1), 90.
78. Robinson, S. A., Reilly, E. D., Petrakis, B. A., Wiener, R. S., Castaneda-Sceppa, C., & Quigley, K. S. (2022). Daily physical activity and sleep in veterans: the role of insomnia severity. *Journal of Behavioral Medicine*, *45*(2), 318-323.
79. Roh, S.-Y., Yeom, H.-A., Lee, M.-A., & Hwang, I. Y. (2014). Mobility of older palliative care patients with advanced cancer: A Korean study. *European Journal of Oncology Nursing*, *18*(6), 613-618.
80. Rosenbaum, S., Vancampfort, D., Tiedemann, A., Stubbs, B., Steel, Z., Ward, P. B., Berle, D., & Sherrington, C. (2016). Among inpatients, posttraumatic stress disorder symptom severity is negatively associated with time spent walking. *Journal of Nervous and Mental Disease*, *204*(1), 15-19.
81. Saraiva, M., Castro, M. A., & Vilas-Boas, J. P. (2022). The Role of Sleep Quality and Physical Activity Level on Gait Speed and Brain Hemodynamics Changes in Young Adults-A Dual-Task Study. *Eur J Investig Health Psychol Educ*, *12*(11), 1673-1681.
82. Serrano-Checa, R., Hita-Contreras, F., Jimenez-Garcia, J. D., Achalandabaso-Ochoa, A., Aibar-Almazan, A., & Martinez-Amat, A. (2020). Sleep Quality, Anxiety, and Depression Are Associated with Fall Risk Factors in Older Women. *International Journal of Environmental Research & Public Health [Electronic Resource]*, *17*(11), 05.
83. Shibata, S., Tsutou, A., & Shiotani, H. (2013). Relation between sleep quality and daily physical activity in hemodialysis outpatients. *Kobe Journal of Medical Sciences*, *59*(5), E161-E166.
84. Shih, Y. H., Wang, J. J., Shih, Y. H., Pai, M. C., & Huang, Y. C. (2017). Sundown Syndrome, Sleep Quality, and Walking Among Community-Dwelling People With Alzheimer Disease. *Journal of the American Medical Directors Association*, *18*(5), 396-401.
85. Siengsukon, C. F., Aldughmi, M., Kahya, M., Lynch, S., Bruce, J., Glusman, M., Ness Norouzinia, A., & Billinger, S. (2018). Individuals with mild MS with poor sleep quality have impaired visuospatial memory and lower perceived functional abilities. *Disability & Health Journal*, *11*(1), 116-121.
86. Silva, H., Mantoani, L. C., Aguiar, W. F., Goncalves, A. F. L., da Silva, T. G., Zamboti, C. L., Ribeiro, M., Probst, V. S., Pitta, F., & Camillo, C. A. (2023). The impact of sleep duration on physical activity in daily life in patients with idiopathic pulmonary fibrosis. *Physiotherapy Theory & Practice*, 1-10.
87. Silva, L. C., Silva, A., Rangel, M. F. A., Caetano, L. C. G., Teixeira-Salmela, L. F., & Scianni, A. A. (2021). Depressive symptoms and functional status are associated with sleep quality after stroke. *Topics in Stroke Rehabilitation*, *28*(8), 573-580.
88. Spira, A. P., Covinsky, K., Rebok, G. W., Punjabi, N. M., Stone, K. L., Hillier, T. A., Ensrud, K. E., & Yaffe, K. (2012). Poor sleep quality and functional decline in older women. *Journal of the American Geriatrics Society*, *60*(6), 1092-1098.
89. Stack, E. L., & Ashburn, A. M. (2006). Impaired bed mobility and disordered sleep in Parkinson's disease. *Movement Disorders*, *21*(9), 1340-1342.
90. Stenholm, S., Kronholm, E., Bandinelli, S., Guralnik, J. M., & Ferrucci, L. (2011). Self-reported sleep duration and time in bed as predictors of physical function decline: results from the InCHIANTI study. *Sleep*, *34*(11), 1583-1593.

91. Stenholm, S., Kronholm, E., Sainio, P., Borodulin, K., Era, P., Fogelholm, M., Partonen, T., Porkka-Heiskanen, T., & Koskinen, S. (2010). Sleep-related factors and mobility in older men and women. *J Gerontol A Biol Sci Med Sci*, *65*(6), 649-657.
92. Sullivan Bisson, A. N., Robinson, S. A., & Lachman, M. E. (2019). Walk to a better night of sleep: testing the relationship between physical activity and sleep. *Sleep Health*, *5*(5), 487-494.
93. Suri, S. V., Batterham, A. M., Ells, L., Danjoux, G., & Atkinson, G. (2015). Cross-sectional Association between Walking Pace and Sleep-disordered Breathing. *International Journal of Sports Medicine*, *36*(10), 843-847.
94. Takemura, N., Cheung, D. S. T., Fong, D. Y. T., Lee, A. W. M., Lam, T. C., Ho, J. C., Kam, T. Y., Chik, J. Y. K., & Lin, C. C. (2021). Relationship of subjective and objective sleep measures with physical performance in advanced-stage lung cancer patients. *Scientific Reports*, *11*(1), 17208.
95. Teas, E., & Friedman, E. (2021). Sleep and functional capacity in adults: Cross-sectional associations among self-report and objective assessments. *Sleep Health*, *7*(2), 198-204.
96. Theodorou, V., Karetsi, E., Daniil, Z., Gourgoulianis, K. I., & Stavrou, V. T. (2020). Physical Activity and Quality of Sleep in Patients with End-Stage Renal Disease on Hemodialysis: A Preliminary Report. *Sleep Disorders*, *2020*.
97. Tighe, C. A., Brindle, R. C., Stahl, S. T., Wallace, M. L., Bramoweth, A. D., Forman, D. E., & Buysse, D. J. (2021). Multidimensional Sleep Health and Physical Functioning in Older Adults. *Gerontology & Geriatric Medicine*, *7*, 23337214211016222.
98. Tyagi, S., Perera, S., & Brach, J. (2015). Balance and Mobility in Community-Dwelling Older Adults: Impact of Daytime Sleepiness. *Journal of the American Geriatrics Society*, *63*, S62-S62. <Go to ISI>://WOS:000352578900173
99. Umemura, G. S., Makhoul, M. P., Torriani-Pasin, C., & Forner-Cordero, A. (2022). Circadian parameter as a possible indicator of gait performance and daily activity levels in chronic stroke survivors. *Annual International Conference Of The IEEE Engineering In Medicine And Biology Society*, *2022*, 4370-4373.
100. Umemura, G. S., Pinho, J. P., Duysens, J., Krebs, H. I., & Forner-Cordero, A. (2021). Sleep deprivation affects gait control. *Scientific Reports*, *11*(1), 21104.
101. Vardar-Yagli, N., Saglam, M., Savci, S., Inal-Ince, D., Calik-Kutukcu, E., Arikan, H., & Coplu, L. (2015). Impact of sleep quality on functional capacity, peripheral muscle strength and quality of life in patients with chronic obstructive pulmonary disease. *Expert Review of Respiratory Medicine*, *9*(2), 233-239.
102. Vaz Fragoso, C. A., Miller, M. E., Fielding, R. A., King, A. C., Kritchevsky, S. B., McDermott, M. M., Myers, V., Newman, A. B., Pahor, M., & Gill, T. M. (2014). Sleep-wake disturbances in sedentary community-dwelling elderly adults with functional limitations. *Journal of the American Geriatrics Society*, *62*(6), 1064-1072.
103. Wang, L., & Zou, B. (2022). The Association Between Gait Speed and Sleep Problems Among Chinese Adults Aged 50 and Greater. *Frontiers in Neuroscience*, *16*, 855955.
104. Xue, F., Wang, F. Y., Mao, C. J., Guo, S. P., Chen, J., Li, J., Wang, Q. J., Bei, H. Z., Yu, Q., & Liu, C. F. (2018). Analysis of nocturnal hypokinesia and sleep quality in Parkinson's disease. *Journal of Clinical Neuroscience*, *54*, 96-101.

105. Yeom, H.-A., Baldwin, C. M., Lee, M.-A., & Kim, S.-J. (2015). Factors Affecting Mobility in Community-dwelling Older Koreans with Chronic Illnesses. *Asian Nursing Research*, *9*(1), 7-13.
106. Zhan, Q., Zhao, J., Guo, Q., Niu, J., Yu, C., Ding, W., Zhang, L., Qi, H., & Shao, X. (2023). Association of Sleep Duration with Physical Performance in Hemodialysis Patients: A Multicenter Cross-Sectional Study. *Nephron*, *147*(5), 260-265.
107. Zhang, L., Liu, S., Li, Y., Li, S., & Wu, Y. (2022). Associations of Sleep Quality with Gait Speed and Falls in Older Adults: The Mediating Effect of Muscle Strength and the Gender Difference. *Gerontology*, *68*(1), 1-7.

| First Author Last Name | Year | Study Aim | Major Sleep-Mobility Results |
|---|---|---|---|
| Ahern | 2019 | Examine the relationship between sleep duration and physical function in non-elderly individuals with severe obesity. | Physical function (speed ascending/descending a step, completion of 500m walk) was better in non-elderly subjects with severe obesity who sleep 6-9 hours compared to their counterparts who reported shorter or longer sleep duration; People with abnormal sleep duration have decreased ability to perform and/or learn a functional motor task. |
| Auyeung | 2015 | Examine how seep could affect testosterone and whether sleep duration and sleep disturbances were associated with testosterone level, muscle mass, grip strength, and walking speed in a group of men older than 65 years of age. | Insomnia was associated with a slower walking speed; Walking speed not associated with go-to bed time, wake-up time or sleep duration. |
| Awotidebe | 2017 | Investigate the relationship between sleep quality and functional capacity in Nigerian patients with Chronic Heart Failure and healthy controls. | Pittsburgh Sleep Quality Index total score, and all subscores except sleep duration had an inverse significant correlation (-0.362, 0.001) with functional capacity in patients with Chronic Heart Failure; Pittsburgh Sleep Quality Index total score had a positive significant correlation with functional capacity; Inverse significant relationship between functional capacity and Sleep Quality (-0.424, 0.001) in patients with Chronic Heart Failure |
| Ayaki | 2014 | Evaluate cataract surgical cases implanted with clear, UV-Blocking Intraocular Lenses by measuring quality of life, sleep quality, and gait speed. | Preoperative gait speed was not associated with change in Pittsburgh Sleep Quality Index ("change" is from preop to 2 months; $p=0.21$); Preoperative Pittsburgh Sleep Quality Index score was not associated with change in gait speed ($p = 0.72$); Change in Pittsburgh Sleep Quality Index was associated with change in gait speed ($p < 0.05$). |
| Batalla-Martin | 2020 | Establish the prevalence of insomnia using the diagnostic criteria of the10th revision of the International Classification of Diseases and the 4th edition of the Diagnostic and Statistical Manual of Mental Disorders to determine its relationship with Health-Related Quality of Life in patients with diagnosed schizophrenia. | The likelihood of insomnia when there are problems in the quality of life is significant in all its dimensions: Mobility Odds Ratio: 3.54 (95% Confidence Interval 1.88-6.65); The presence of insomnia increases the likelihood of having problems with mobility. |
| Benito-Villena | 2022 | Examine the impact of regular physical activity on the prevention of insomnia in pregnant Spanish women. | At 19th Gestational Week, groups I1 and I2 reached a mean of 6267 steps/day (Standard Deviation = 3854) and 5835 steps/day (Standard Deviation = 2741), respectively ($p > 0.05$); At 31st Gestational Week, mean steps/day was lower for I2 ($p < 0.001$); Insomnia and poor sleep quality prevalence increased through pregnancy, but no differences between groups were found ($p > 0.05$); Lineal regression showed no association between the average steps/day at third trimester of pregnancy and Athens Insomnia Scale and Pittsburgh Sleep Quality Index scores; The walking promotion program based on pedometers did not help to prevent insomnia in the third trimester of pregnancy. |
| Bernstein | 2019 | Expand the literature on the relationship between sleep and gait by examining relationships between different aspects of gait and 3 different measures of self-reported sleep in a large, cognitively healthy sample of older adults. | Poorer sleep quality was associated with greater Single Task asymmetry ($F[8, 493] = 24.33$, $t[501] = -2.57$, $\beta = 0.16$, $p < 0.05$); Poorer sleep quality was also associated with greater Dual Task asymmetry ($F[8, 493] = 14.76$, $t[501] = -2.62$, $\beta = 0.17$, $p < 0.01$); Greater daytime sleepiness was associated with increased Dual Task pace variability/postural control ($F[8, 493] = 3.23$, $t[501] = 2.53$, $\beta = 0.12$, $p < 0.05$). |
| Bertapelli | 2022 | Examine sleep quality indicators and its association with physical functioning in adults with Down Syndrome, and whether associations are altered by Body Mass Index and age. | Total sleep time was significantly associated with 6-Meter Walking distance; The average length of awakenings was negatively associated with 6-Meter Walking distance; Sleep latency was moderately associated with 6-Meter Walking distance; Sleep efficiency showed a moderate association with 6-Meter Walking distance; Total time in bed was significantly associated with Timed Up and Go performance after controlling for age and Body Mass Index. |
| Boolani | 2022 | To use machine learning algorithms to identify individuals who report Acute Partial Sleep Deprivation, Sleep Extension, and those who report 7-9 hours of sleep. | Those who obtain 7-9 h of sleep the prior night exhibit a different single-task gait pattern than those who slept less or more; Those who self-reported 7-9 h of sleep had more asymmetrical gait with slower gait speed and less trunk motion as compared to those who reported acute sleep deprivation or sleep extension; Those who reported Acute Partial Sleep Deprivation or sleep extension had trouble maintaining gait speed as evidenced by increased variance in cadence and larger stride lengths and less time spent in single leg support time as compared to those who received the recommended amount of sleep; The results suggest getting more or less than the recommended amount of sleep had gait patterns consistent with those who are trying to ambulate faster but are unable to maintain a steady gait speed as evidenced by increased variance in cadence, gait speed, and circumduction by individuals who report more or less than the recommended amount of sleep; An interesting finding in our data was the similarities in gait between those who reported sleep deprivation compared to those who reported sleep extension. |
| Chasens | 2012 | Examine the relationship between physical activity and symptoms of insomnia among adults with prediabetes. | Men walked more steps than women; however, women had more insomnia symptoms; There were significant associations between insomnia symptoms and increased sleep latency and decreased sleep duration; Multiple regression analysis showed that fewer insomnia symptoms were significantly related to increased steps walked. |
| Chen | 2022 | Examine the association between daytime physical activity, while-in-bed smartphone use, sleep delay, and sleep quality. | Participants who claimed smartphone-related sleep delay generally had fewer walking steps; We found associations between walking steps and while-in-bed smartphone use and sleep delay, but the estimates were subtle (Odds Ratio = 1.000)" |

| | | | |
|---|---|---|---|
| Chen | 2018 | Examine the independent relationships between frequency, duration, intensity of walking, and walking volume with self-reported sleep difficulty in older adults 2 years later. | Participants with low walking volume (p < 0.001), low duration (p = 0.001), and low speed (p = 0.023) at baseline had an increased sleep difficulty during this period when compared to the high-level groups; Association between walking frequency and sleep-score changes was only marginally significant (p = 0.064) and walking duration as a significant predictor (p = 0.012) among 3 walking parameters. |
| Cheng | 2022 | Investigate the relationship between walking activity (frequency, intensity, and duration), and sleep quality. | Total weekly walking time (P = .000), walking days per week (P = .019), walking frequency per day (P = .002), and time spent (minutes) per walk (P = .018), showed significant correlation with sleep quality; Total weekly walking time and frequency of walking per day were significant predictors of sleep quality; For those who walked for less than 210 minutes a week, the likelihood of having poor sleep quality was 2.7 times higher than those who walked for more than 210 minutes per week (odds ratio: 2.700, 95% confidence interval: 1.388–5.250); For those who walked once per day, the likelihood of having poor sleep quality was 2.2 times higher than for those who walked more than once per day Odds Ratio: 2.198, 95% Confidence Interval: 1.013–4.770) |
| Chevance | 2022 | Estimate whether physical activity on one day was associated with both sleep quality and quantity the following night; Determine what extent sleep on one night was associated with physical activity the next day. | Results suggest an absence of association between steps and sleep efficiency in the two directions for most participants; Association between steps and total sleep time, with 58% of our participants showing a negative association between total sleep time and next day steps, and 27% showing a negative association between steps and next day total sleep time; The association between steps and total sleep time was significant in both directions for 5 participants, thus indicating bi-directional associations; 4 participants presented a significant and negative association between steps and total sleep time in the two directions, more steps on one day was associated with shorter sleep duration the following night, and shorter sleep duration on one night was associated with more steps the following day; One participant presented a singular pattern of results: more steps on one day was associated with longer sleep duration the following night, but, as for the other participants, shorter sleep duration on one night was associated with more steps the following day. |
| Chien | 2015 | Examine if the relationship between poor sleep quality and physical disability in older adults | Older adults with poor sleep quality were found to have a higher likelihood of suffering from a physical disability (Odds Ratio: 2.03; 95% Confidence Interval: 1.02–4.05; p = 0.04); |
| Cole | 2009 | Examine relationships among disordered sleep and cognitive and functional status in nursing home residents. | Decreased Total Sleep Time was associated with better functional status measured as gait speed (r = 0.32, p=0.01); Sleep efficiency, Sleep Onset Latency, awakenings, and Wake After Sleep Onset were not significantly associated with gait speed; After controlling for the effect of cognitive status, the association between better gait speed and decreased Total Sleep Time remained significant (r = 0.24, p=0.03) |
| Curran | 2022 | Evaluate physical activity and sedentary behavior levels in people with cystic fibrosis and determine their association with aerobic capacity, lung function, and sleep. | Poor sleep quality was strongly and significantly negatively correlated with low step counts (r = -0.85, P < .001) |
| Dam | 2008 | Determine whether sleep quality is associated with physical function in older men. | Decreased nighttime sleep was associated with walking speed, and increased odds of not completing the chair stand in age-adjusted analyses; Men with the least amount of Rapid Eye Movement sleep (<14.8%) had the slowest walking speed; No consistent associations were seen with stage 1, stage 2 or stage 3/4 sleep and physical function; Men with sleep apnea (Respiratory Disturbance Index > 30) had 5.2% slower walking speed than men without sleep apnea after adjustment for age Men with Respiratory Disturbance Index > 30 had approximately twice the odds of not completing one chair stand; After adjusting for covariates, sleep apnea was not significantly associated with any of the four physical function measures; Analyses substituting Polysomnography measures for Total Sleep Time, Sleep Efficiency, Sleep Latency and Wake After Sleep Onset were consistent with the actigraphy results for the walking speed , but not for inability to do chair stand. |
| Davis | 2019 | Examine the association of subjective sleep parameters with depression, health related quality of life, physical function, and cognition among stroke survivors. | Physical function and health was significantly associated with Pittsburgh Sleep Quality Index-subjective sleep quality, Pittsburgh Sleep Quality Index-sleep latency, Pittsburgh Sleep Quality Index-sleep duration, and Pittsburgh Sleep Quality Index-daytime dysfunction; Multivariate linear regression demonstrated that Pittsburgh Sleep Quality Index-daytime dysfunction predicted physical function; Sleep duration (b = .0014 (.0007); P= .054); $r^2$ = .05) was associated with physical function (meters walked in 6 minutes). Habitual sleep efficiency (b = .04 (.01); P= .002; $r^2$ = .13) was associated with physical function (i.e. meters walked in 6 minutes); One component, daytime dysfunction, of the Pittsburgh Sleep Quality Index was associated with physical function (meters walked in 6 minutes) (b = -66.6 (22.4); P= .004, $r^2$ = .15) after adjusting for age, sex, and cognitive variables. |
| Denison | 2021 | Examine the association between sleep quality and physical performance among a group of community-dwelling older adults, according to sex. | Poor sleep quality (Pittsburgh Sleep Quality Index >5) was not associated with 8-ft walk time, chair rise test, Timed Up and Go time, or tandem stand performance among men or women; In men, more sleep disturbance was associated with a higher likelihood of a poor score on the short physical performance battery; In women, poor sleep was associated with a lower likelihood of a poor score on the short physical performance battery. |
| Diaz-Abad | 2018 | To determine whether poor sleep quality was present in patients with newly diagnosed Amyotrophic Lateral Sclerosis and whether risk factors associated with poor sleep quality were present. | There was no difference in Pittsburgh Sleep Quality Index based on the Amyotrophic Lateral Sclerosis Functional Rating Scale at different severity cutoffs; Although sleep quality was not associated with functional status as assessed by the Amyotrophic Lateral Sclerosis Functional Rating Scale score, the patient's ability to turn in bed and adjust bed clothes was associated with poor sleep quality, r = -0.335 (P = - 0.033). |

| | | | |
|---|---|---|---|
| Dobrosielski | 2016 | Examine the relationship between vasodilatory capacity, markers of physical activity, and self-reported sleep quality in a group of young and older community-dwelling adults. | Total Sleep Time was negatively associated with steps/day (young r = -0.31, p = 0.02; old r = -0.47, p<0.01; combined sample r = -0.39, p<0.01; Wake After Sleep Onset and Pittsburgh Sleep Quality Index were not significantly associated with steps/day. |
| Edgar | 2021 | Investigate the relationship between sleep, physical performance, and the subjective wellbeing of officer trainees during 6 weeks of initial military training. | Sleeping more than 6:15 (hr:min) per night over 6 weeks was associated with small benefits to aspects of physical performance, and moderate to large benefits on subjective wellbeing measures when compared with sleeping < 6:15 hr:min; Moderate relationships were observed between time in bed and faster 2.4-km run time (r= -.47); for sleep-onset latency and press-ups (r= .48), and for wake after sleep onset and press-ups (r= -.47); Only weak or very weak relationships were found for all other measures. |
| Endeshaw | 2009 | Identify the risk factors associated with the onset and progression of cardiovascular disease. | A sleep-related complaint was more likely to be reported by those who had a slow walking speed compared with those with normal walking speed (26% vs. 16%, respectively; x2 =9.66, P =0.001); The relation between severe Sleep Disordered Breathing and slow walking speed was statistically significant among women (X2 = 12.33, P < 0.001), but not among men. |
| Ezeugwu | 2017 | Objectively describe whole-day activities (including sleep) within 1 month after inpatient stroke rehabilitation; Explore the relationships between accelerometer-derived variables with age, time since stroke, gait speed, cognitive scores, and quality-of-life ratings. | A higher sleep duration was significantly correlated with less time standing, less number of sit-to-stand transitions, and a lower number of purposeful steps. Sleep duration and gait speed significantly correlated at -.17, steps at -.24, sit-to-stand transitions at -.51. |
| Feng | 2018 | Examine the level of physical activity in adults with untreated moderate-to-severe obstructive sleep apnea with obesity versus those without obesity; Investigate the change in physical activity following 4 months of positive airway pressure treatment. | Following 4 months of positive airway pressure treatment, participants without obesity and with Obstructive Sleep Apnea showed significant improvements (763.5 (25.7, 1501.3); p = 0.043) in steps per day on waist accelerometer. |
| Fex | 2012 | Examine the relationship between long sleep duration and functional capacity in sedentary post-menopausal women. | Significant negative correlation between sleep hours and chair stand test (r = -.033, p = 0.02); Sleep duration was the primary independent predictor of the chair stand test explaining 10.7% of the variance; There were no difference between number of steps per day between normal and long sleepers (p = 0.7). |
| Fleming | 2020 | Assess the relationship between sleep quality and motor recovery in brain injury patients receiving inpatient rehabilitation. | Higher functional independence at admission and less disrupted sleep over the rehabilitation period was associated with better mobility at discharge. Wake After Sleep Onset did not contribute to the model. If a stepwise regression was used, sleep fragmentation explained significant variance in Rivermead Mobility Index (R2=0.324, P<.001). |
| Fongen | 2024 | Examine the prevalence of reduced sleep quality and to identify aspects of sleep that are affected in a group of patients with axial spondyloarthritis with moderate-to-high disease activity; Assess the association between sleep quality and physical function, cardiorespiratory fitness, and spinal mobility. | Patients with normal sleep quality had statistically significantly better performance-based physical function (28.9 ± 7.2 seconds) than patients with reduced sleep quality (34.0 ± 11.4 seconds); Adjusted for age and sex, for each second less needed<br>in Ankylosing Spondylitis Performance Index, the Pittsburgh Sleep Quality Index score was reduced (Î$^2$ = 0.10, 95% Confidence Interval 0.01, 0.19). |
| Fritschi | 2017 | Examine differences in physical function between older and younger adults with Type 2 Diabetes and when these changes occur; Compare differences in predictors of physical function between the older and younger cohorts. | Bivariate analyses of factors related to physical function as measured by 6-Minute Walking Distance showed significant associations (all p < 0.05) with sleep quality (Wake After Sleep Onset, r = -0.302). |
| Fulk | 2020 | Evaluate the impact of self-reported sleep problems on recovery of health-related quality of life and participation in people with stroke. | At 2 months post stroke, there were group differences in mobility subscale, with post-hoc analyses determining the difference was between the no sleep problem group and the moderate-to-quite-a-bit sleep problem group; At 6 months post stroke, there were group differences in the mobility subscale, with post-hoc analyses determining a difference between the no sleep problem and moderate-to-quite-a-bit sleep problem group as well as difference between the no-to-minimal sleep problem and moderate-to-quite-a-bit sleep problem group; At 12 months post stroke, there were group differences in the mobility subscale, with post-hoc analyses determining a group difference between the no sleep problem and moderate-to-quite-a-bit sleep problem group as well as difference between the no-to-minimal sleep problem and moderate-to-quite-a-bit sleep problem group. |
| Goldman | 2007 | Examine the association of objective measures of sleep and nap patterns with objective measures of neuromuscular performance and subjective measures of daytime function. | In the unadjusted model for nighttime sleep duration and gait speed women who averaged <6 hours and ≥7.5 hours of Total Sleep Time performed more poorly on the 6-meter usual pace test (meters/sec); In the fully adjusted model for gait speed, women who averaged <6 hours sleep per night still had 3.5% slower gait speed than the women who averaged 6-6.8 hours of sleep; Nighttime sleep duration was significantly associated with time to complete 5 chair stands in both the adjusted and |

| | | | |
|---|---|---|---|
| | | | unadjusted models. In the fully adjusted model for chair stand time, women who averaged >7.5 hours of sleep per night took 4.1% longer to complete 5 chair stands than those who slept 6.8-7.5 hours/night. |
| Gonzalez-Sanchez | 2019 | Evaluate the associations between insomnia, physical activity, and diet quality in an adult population. | The mean differences of total steps/day, aerobic steps/day, and kilocalories/day spent in performing physical activity, between no insomnia group and insomnia group were:: 1022.5 (95% CI:177.9–1867.0), 743.9 (95% CI: 68.3–1419.4) and 39.8 (95% CI: 5.7–73.9), respectively; The findings suggest that completing daily a greater total number of steps, aerobic steps, and energy expended by walking, could be correlated with less insomnia, independent of age, sex, and other confounding variables. |
| Greysen | 2022 | Examine the relationship between daily sleep and physical activity with 6 patient-reported functional outcomes (symptom burden, sleep quality, physical health, life space mobility, activities of daily living, and instrumental activities of daily living) at 13 weeks after hospital discharge. | Participants who had both adequate sleep (7-9hours/night) and activity (>5000 steps/day) had better functional outcomes at 13 weeks after hospital discharge. Participants with adequate sleep but less activity (<5000 steps/day) had significantly worse symptom burden (z-score 0.93, 95% Confidence Interval 0.3 to 1.5;P=.02), community mobility (z-score –0.77, 95% Confidence Interval–1.3 to –0.15; P=.02), and perceived physical health (z-score –0.73, 95% Confidence Interval –1.3 to –0.13; P=.003), compared with those who were more physically active (≥5000 steps/day). |
| Gururaj | 2021 | Assess the relationship between physical activity, sleep, and circadian rhythm using accelerometer and urine melatonin levels in patients with head and neck cancer during the course of chemoradiotherapy. | Sleep efficiency at baseline was significantly negatively correlated with step count (r = -0.65, p < .0001). Sleep efficiency at 3-weeks of chemoradiation was significantly negatively correlated with step count (r = -0.31, p = .15). At the end of 7 weeks, Total Sleep Time was found to be moderately correlated to step count (r = 0.35, p = .12). |
| Hellstrom | 2023 | Investigate whether symptoms of insomnia are associated with self-reported health and life satisfaction in cardiac arrest survivors 6 months after the event. | 20% of the sample reported clinical insomnia; Insomnia was significantly associated with all aspects of self-reported health (p < 0.01) and life satisfaction (p < 0.001), except mobility (p = 0.093), self-care (p = 0.676), and usual activities (p = 0.073). |
| Hirata | 2020 | Evaluate whether time spent in physical activity and in sedentary behavior are associated with sleep quantity and quality in individuals with Chronic Obstructive Pulmonary Disease. | There's a significant decrease in 6-Minute Walk Test Distance as Time in Bed increases, indicating that longer time in bed is associated with lower walking distance (p-value: 0.001); An increase in Total Sleep Time is associated with a significant decrease in 6-Minute Walk Test Distance (p-value: 0.001); Lower sleep efficiency is associated with a decrease in 6-Minute Walk Test Distance (p-value: 0.025); More sleeping bouts are associated with a decrease in 6-Minute Walk Test distance (p-value: 0.024); No significant correlation with 6-Minute Walk Test distance (p-value: 0.429); Increased Wake After Sleep Onset is associated with a decrease in 6-Minute Walk Test distance (p-value: 0.006); A significant decrease in steps per day is observed with an increase in Time in Bed (p-value: <0.0001); An increase in Total Sleep Time is associated with a significant decrease in steps per day (p-value: <0.0001); Lower sleep efficiency is associated with fewer steps per day (p-value: <0.025); Increased Wake After Sleep Onset is associated with fewer steps per day (p-value: 0.006). |
| Holingue | 2024 | Determine the association of actigraph-measured sleep with self-report and objective measures of physical function among community-dwelling older men. | Participants with short average baseline total sleep time (< 6 hours) had significantly greater slowing in their walking speed from baseline to follow-up; Participants with long baseline sleep onset latency (>30 minutes) had significant increases in mobility difficulties and time to complete chair stands; Sleep efficiency and wake after sleep onset were not significantly associated with any outcomes; No sleep predictors were associated with change in instrumental activities of daily living. |
| Howell | 2018 | Examine how self-reported sleep duration during the night prior to baseline concussion testing affects single-task and dual-task abilities during an instrumented gait examination and a timed tandem gait test. | Steady-state gait measurements were not significantly different between sleep groups; Differences between sleep time groups were observed for the best time (main effect of group: p = .001; Î·p2 = 0.06) and mean time (p < .001; Î·p2 = 0.07) for both single-task and dual-task tandem gait conditions, where those who slept for > 7h the night prior to tested completed the tandem gait faster than those who slept for less than 7h during the night prior to testing. |
| Huang | 2021 | Investigate the relationships between accelerometer-measured sleep duration and physical function in a younger-old Taiwanese population after adjusting for objectively measured physical activity. | A positive association of sleep duration with grip strength was found after adjusting for covariates (P = .005); No significant associations were observed between sleep duration and the other physical function outcomes. |
| Hur | 2022 | Determine the association between sleep measurements by wrist actigraphy and patient self-report; Assess the impact of physical activity and extrapulmonary symptoms on sleep quality in patients with Fibrotic Interstitial Lung Disease. | Total sleep times from the Pittsburgh Sleep Quality Index and wrist monitors showed weak (r ¼ -0.20, p ¼ 0.04) and moderate (r ¼ -0.38, p < 0.001) correlations with daily step count, respectively; Self-reported and actigraphy measurements of sleep efficiency, sleep latency, and number of sleep disturbances were poorly correlated with physical activity; Daily step count did not predict sleep quality when adjusting for covariates, with exception of a statistically significant adjusted association between greater daily step count and shorter total sleep time measured by the wrist monitor. |
| Izawa | 2011 | Determine differences in physical activity and health-related quality of life relating to sleep quality; Determine target values of physical activity that would improve sleep quality and health-related quality of life in outpatients with chronic heart failure. | After adjustment for New York Heart Association class, step counts in the Shallow Sleep group were found to be significantly lower than those of the Deep Sleep group (5541.5 [296.5] steps/day for 1 week, 95% Confidence Interval: 4957.1 - 6129.8 vs. 6811.4 [306.7] steps/day for 1 week, 95% Confidence Interval: 6208.8 - 7421.3, F = 8.87, p = 0.003); After adjustment for New York Heart Association class, the mean Epworth Sleepiness Scale score in the Shallow Sleep group was significantly higher than that of the Deep Sleep group (8.05 [4.14] points, 95% Confidence Interval: 7.19 -8.91 vs. 5.45 [0.44] points, 95% Confidence Interval: 4.57 - 6.34, F = 17.30, p < 0.001). |

| | | | |
|---|---|---|---|
| Jean | 2017 | Evaluate correlation between changes in physical activity and improvements in sleep parameters in patients with obstructive sleep apnea after treatment with Continuous Positive Airway Pressure (CPAP). | Before Continuous Positive Airway Pressure (CPAP) treatment, lower steps/day was seen in patient with poor sleep quality (r = -0.36, p=0.004); Steps/day increased after CPAP use (F(1.46, 86)=44.6, p<0.001); Post hoc tests using the Bonferroni correction revealed that CPAP use elicited a significant improvement in pedometer steps from pretreatment/baseline to visit 1 after initiation of CPAP use (P = <0.001); Furthermore, at the second visit post-treatment, there was a further increase in pedometer steps, compared with pretreatment/baseline (P < .001) and visit 1 (P < .001). |
| Jeon | 2016 | Evaluate the extent to which sleep-related daytime symptoms (fatigue, sleepiness, and depressive symptoms) mediate the relationship between sleep disturbance and functional performance among patients with stable heart failure. | Only sleep quality was associated with 6-Minute Walk Test (r=-0.22, p-value not provided). There were no statistically significant, indirect effects on objective physical function (6-Minute Walk Test) through the daytime symptoms. |
| Kaizu | 2022 | Clarify the distribution of physical activities, including sleep, sedentary behavior, low intensity physical activity, and moderate-to vigorous physical activity of hospitalized older adults with trunk and lower extremity fractures; Clarify the details of the intensity of each physical activity and the relationship between physical activity and physical function. | Each of the physical activity intensity levels (including sleep) and the Timed Up and Go test results were not significantly correlated |
| Kasovic | 2021 | Examine the associations between gait speed and sleep quality in first-year university students, according to gender. | In the unadjusted model, faster participants had significantly “better” sleep quality (β=−3.15, 95% CI −3.82 to −2.47, p<0.001). When the model was adjusted for sex, age, body-mass index, self-rated health, smoking status, and psychological distress, faster participants remained having “better” sleep quality (β=−2.88, 95% CI −3.53 to −2.22, p<0.001); “Slower” gait speed was correlated with “poorer” subjective sleep quality (r=−0.36, p<0.001), “longer” sleep latency (r=−0.40, p<0.001), “shorter” sleep duration (r=−0.58, p<0.001), “poorer” sleep efficiency (r=−0.34, p<0.001), “having more” sleep disturbances (r=−0.40, p<0.001), “using” sleeping medications (r=−0.21, p<0.001) and “greater” daytime dysfunction (r=−0.60, p<0.001). |
| Kataoka | 2020 | Evaluate the associations between objective sleep measures and morning mobility among patients with Parkinson's Disease. | Univariable linear regression models demonstrated significant associations between objective sleep measures and morning mobility (Sleep Efficiency per %: β, −0.285; 95% Confidence Interval = −0.560 to −0.010; p = 0.042; $R^2$ = 0.026; Wake After Sleep Onset per min: β, 0.101; 95% Confidence Interval = 0.037 to 0.165; p = 0.002; $R^2$= 0.059; and FI per unit; β, 1.245; 95% Confidence Interval = 0.140 to 2.350; p = 0.028; $R^2$ = 0.031); A multivariable linear regression model fully adjusted for potential confounders still demonstrated significant associations between better sleep measures and shorter time in low mobility below 100 counts/min within 2 hours after getting out of bed (Sleep Efficiency per %: β, −0.419; 95% Confidence Interval = −0.635 to −0.204; p < 0.001; Wake After Sleep Onset per min: β, 0.056; 95% Confidence Interval = 0.003 to 0.109; p = 0.039; and FI per unit; β, 1.161; 95% Confidence Interval = 0.300 to 2.023; p = 0.009) but not TST (p = 0.78). These associations were of similar strength using 50 counts/min within 1 hour after getting out of bed as the low-mobility threshold. |
| Kaya | 2023 | Identify the relationship between sleep quality and gait speed in geriatric patients. | A positive correlation was found between the total Pittsburgh Sleep Quality Index score and gait speed time (r=0.250, p=0.003); Poor sleepers had significantly slower gait speed than good sleepers; ROC analysis revealed that a 4-m gait speed time significantly predicted poor sleep quality in geriatric patients. |
| Kim | 2015 | Examine the association between objective measures of sleep quality and performance-based measures of physical function in community-dwelling Japanese adults ≥80 years. | Sleep efficiency and wake after sleep onset were independently associated with maximum walking speed (β = 0.277; 95% CI 0.103 to 0.351, P < 0.001 and β = −0.214; 95% Confidence Interval −0.339 to−0.082, P = 0.001, respectively); Sleep efficiency and wake after sleep onset was independently associated with usual walking speed (β = 0.200, 95% Confidence Interval 0.035 to 0.305, P = 0.005 and β = −0.174; 95% Confidence Interval−0.341 to −0.064, P = 0.004, respectively); Wake after sleep onset was not associated with handgrip strength; There were no associations between physical function and total sleep times less than 6 h or greater than 8 h, and number of awakenings in all models. |
| Kim | 2019 | Identify if the demographic, health-related and stroke-related characteristics of stroke survivors are associated with sleep time; Identify differences in each area of health-related quality of life by sleep time in stroke survivors; Determine the relationship between sleep time and quality of life among stroke survivors. | An association between mobility and sleep was reported. A significant difference was found between sleep duration groups (p=0.011); Participants with ≤5 hours of sleep reported the highest mobility issues (57.3%), followed by those with ≥9 hours (61.1%), while those with 6-8 hours had the lowest (41.6%) |
| Kline | 2021 | Examine the association between trajectories of self-reported insomnia symptoms and sleep duration over a 13-year span with objective physical function assessed at the end of the midlife period (using data from the Study of Women's Health | Compared to those in the "persistent insufficient" sleep duration trajectory group, women in the "persistent sufficient" group had significantly faster times for the 40-ft walk, 4-m walk, and repeated chair stand tests. But this difference did not hold in the minimally or fully adjust models; Following basic covariate adjustment, the consistently low insomnia symptom group had a significantly faster 40-ft walk speed than the consistently high insomnia symptom group (4.6% faster; p = .002) and a |

| | | | |
|---|---|---|---|
| | | Across the Nation); Identify correlates of insomnia symptoms and sleep duration during midlife. | significantly faster 4-m walk speed than the improving and severe worsening insomnia symptom groups (4.2% [p = .01] and 4.1% [p = .04] faster, respectively). In the fully adjusted model, associations remained similar but smaller in magnitude (i.e. 40-ft walk speed for the consistently low group was 3.5% [p = .04] faster than the consistently high group, 4-m walk speed for the consistently low group was 3.7% faster [p = .04] than the improving group). The only exception was that the 4-m walk speed for the consistently low group was no longer significantly faster than the severe worsening group following adjustment for multiple comparisons (3.2% faster; p = .18).The consistently low insomnia symptom trajectory group had a significantly faster repeated chair stand time compared to the consistently high group (5.5% faster; p = .03), but that association was no longer significant in Model 2 (3.1% faster; p = .40). |
| Kubala | 2020 | Examine whether physical activity, measured by self-report and objective methods, was related to a individual components and/or a composite measure of sleep health. | No differences in sleep health were observed across categories of steps/day (F2,94 = 0.58, p=.56, n2=0.01); Steps/day was not associated with any individual sleep health dimensions. |
| Kurose | 2019 | Investigate the association of physical activity with sleep quality and cognitive function in elderly patients with cardiovascular disease in the cardiac rehabilitation maintenance phase. | Locomotor activity was significantly associated with sleep latency (r = -0.222, p = 0.025) but not significantly associated with Pittsburgh Sleep Quality Index global score (r = -0.193, p = 0.051) or total sleep time (r= 0.108, p = 0.280); Locomotor activity was an independent predictor of sleep latency (B = -0.213, p = 0.034) after adjustment for age, sex, cardiac function, and hypnotics. |
| Lavoie-Tremblay | 2014 | Describe the impact of a pedometer-based activity program on a subset of nurses in a hospital setting. | The pedometer program assisted nurses with their sleep problems, and the effect persisted for up to 6 months post-intervention. |
| Lee | 2020 | Determine the associations between physical fitness performance and short and long sleep durations among older adults in Taiwan. | Long-duration sleep was associated with lower body endurance (30 second chair stand), lower body flexibility (chair sit-and-reach test), dynamic balance (8-foot up-and-go). |
| Lee | 2017 | Investigate whether slow gait speed is associated with cognitive impairment and if the association is modified by obstructive sleep apnoea. | Obstructive Sleep Apnea caused no difference in the walking time (p = 0.082 at fast gait speed and p = 0.082 at usual gait speed). |
| Lee | 2020 | Investigate gait characteristics in elderly women aged over 65 years in the subthreshold insomnia stage under various conditions, such as slower, preferred, and faster speeds. | The insomnia group indicated lower pace parameters (range of Cohen's d: 0.283–0.499) and the single support phase (Cohen's d:0.237), greater gait variability (range of Cohen's d: 0.217–0.506), and bilateral coordination (range of Cohen's d: 0.254–0.319), compared with their age-matched controls. The insomnia group demonstrated insufficient gait adaptation at the slower and preferred speeds, as indicated by the Coefficient of variance of the stride length, stride time, and step time. |
| Lee | 2023 | Evaluate the association of demographic factors, injury-related factors, and functional outcomes with the following psychological factors observed in persons with spinal cord injuries: depression, anxiety, sleep quality, and suicide risk. | Sleep Quality (Insomnia Severity Index score) was negatively correlated with the mobility/ambulation domain of the Spinal Cord Independence Measure, but not the Walking Index for Spinal Cord Injury II. |
| Lorenz | 2014 | Explore the relationship between sleep as measured by overnight polysomnography and self-reported physical function in community-dwelling adults. | Statistically significant associations between total sleep time and mobility (r =−0.36; P = .011); Regression analysis indicated that total sleep time explained 12.7% of the variance in mobility (P = .010) after controlling for chronic disease burden, with a 1-minute increase in total sleep time resulted in a 0.02 unit decrease in mobility difficulty score. |
| Louter | 2013 | Assess the influence of subjectively impaired bed mobility on both subjective and objective sleep parameters in insomnia Parkinson Disease patients with and without concerns of impaired bed mobility and controls with primary insomnia. | Of the 44 subjects with Parkinson's Disease with insomnia, 24 54% also reported impaired bed mobility with difficulties turning around or finding a comfortable sleep position (Parkinson's Disease +Impaired Bed Mobility); There were no differences in subjective nocturnal sleep quality or daytime sleepiness between Parkinson's Disease + Impaired Bed Mobility vs Parkinson's Disease - Impaired Bed Mobility subjects; The Polysomnography analyses revealed differences in objective sleep parameters. Total Sleep Time and Sleep Efficiency were significantly lower in the Parkinson's Disease + Impaired Bed Mobility group; This finding also was reflected by a higher number of awakenings and a larger percentage of time spent awake at night. Actigraphy showed no difference between activity periods, duration of periods, or activity levels during sleep or wake between Parkinson's Disease + Impaired Bed Mobility & Parkinson's Disease - Impaired Bed Mobility subjects. |
| Lü | 2017 | Determine the effect of a 24-week Tai Ji Quan training program on self-reported sleep quality, physical performance, and quality of life among elderly Chinese women with knee osteoarthritis. | Improvements in sleep quality were not associated with changes in Timed Up and Go (r and p value not provided). |

| | | | |
|---|---|---|---|
| MacCarthy | 2022 | Evaluate correlations between objective gait measurements via instrumented gait analysis (both longitudinally and cross-sectionally) and cross-sectional patient-reported outcome data across domains of physical, emotional, and social function. | Subscores for anxiety, depression, sleep, and fatigue did not significantly correlate with any instrumented gait analysis data; Walking speed correlated strongly with multiple subscores: physical function (p<0.001, $r_s$=0.708); participation in social roles (p=0.007, $r_s$=0.319); executive function (p=0.005, $r_s$=0.335); Pain interference correlated with longitudinal change in adjusted walking speed (p=0.032, r=–0.259). |
| Malinowska | 2016 | Examine the relationship between sleep quality and mobility disorder in community-dwelling older adults. | After adjustment for age, sex, and depression, both those who did not often sleep well and those with poor awakening were more likely to show Mobility Disorder (Odds Ratio 1.45, 95% Confidence Interval 1.29–1.75), (Odds Ratio 1.77, 95% Confidence Interval 1.50–2.08), respectively; When grouped by both questions, those in the bad awakening group had the least favorable outcome with increased odds ratio for possessing a Mobility Disorder (Odds Ratio 1.95, 95% Confidence Interval 1.61–2.37), compared with good awakening group (Odds Ratio 1.40, 95% Confidence Interval 1.18–1.67). |
| Mantovani | 2016 | Identify correlates of daily step counts measured using pedometers; Analyze the associations between health outcomes and 3 proposed step-count guidelines. | Adults who reached ≥7500 steps/day had a lower likelihood of being obese (odds ratio = 0.38, 95% confidence interval, 0.17–0.85) and reporting worse sleep quality (Odds Ratio = 0.58, 95% Confidence Interval, 0.34–0.99). Adults who reached <5,000 steps/day had a higher likelihood of reporting worse sleep quality (Odds Ratio= 2.11, 95% Confidence Interval, 1.17–3.82). |
| Mei | 2022 | Identify the possible influencing factors of health-related quality of life and its domain-specific contents in stroke patients in rural areas in China. | Patients with stroke with good sleep quality reported fewer issues with mobility than those with poor sleep quality. |
| Moon | 2018 | Investigate the impact of sleep disturbance after stroke when adjusting for other factors such as clinical, demographic, and lesion-related variables to predict the amount of functional gain, (especially in balance and gait) after a 4-week rehabilitation program in subacute stroke patients. | Of the 140 patients, 35 had sleep disturbance (25%); Functional Independence Measure, Functional Ambulation Category, 10-m gait velocity, functional reach, and Berg Balance Scale differed significantly between groups both at baseline and at the 1-month follow-up; Patients with sleep disturbance tended to have worse balance and gait function; 10m velocity for no sleep disturbance = .47± (.47) and with sleep disturbance = 0.18± 0.29 (p=0.002); Functional Ambulation Category = 2.55 (1.97) for no sleep disturbance vs. 1.32(1.53) p=0.001 with sleep disturbance. |
| Motohashi | 1999 | Investigate the relationship between sleep-wake rhythm, physical fitness ,and the competence level of elderly stroke survivors living at home and receiving community rehabilitation services, with special attention to a synchronization of social and biological rhythms. | Earlier rising time was associated with shorter 10-meter walking time (r = 0.33, p < .05); Bedtime was not correlated with 10-m walk time; Rising time was not correlated with time to stand up from bed. |
| Nakakubo | 2018 | Examine whether excessive daytime sleepiness is associated with gait parameters such as speed and variability in community-dwelling older adults. | Excessive Daytime Sleepiness was significantly associated with slower gait speed among younger subjects (<75 years, p = .021) and with both slower gait speed (p = .045) and greater variability in stride length among older subjects (>75 years, p = .048) in a multivariate analysis adjusted for age, sex, body mass index, medication, number of comorbidities, and education. Among younger old adults, 12.4% were classified as having Excessive Daytime Sleepiness, in comparison with 19.5% of older old adults (p < .001). Among older old adults, participants with Excessive Daytime Sleepiness were more likely to have a reduced grip strength when compared with participants without Excessive Daytime Sleepiness (p = .023 and .029, respectively). Among younger old adults, participants with Excessive Daytime Sleepiness slept less than those without Excessive Daytime Sleepiness (p < .001), whereas there was no significant difference between those with or without Excessive Daytime Sleepiness among older old adults (p = .462). In a univariate analysis, participants with Excessive Daytime Sleepiness had a slower gait speed than did participants without Excessive Daytime Sleepiness, regardless of age group (younger: p < .001; older: p = .002). Participants with Excessive Daytime Sleepiness among older old adults showed greater stride length coefficient of variation than participants without Excessive Daytime Sleepiness, but the association between Excessive Daytime Sleepiness and stride length coefficient of variation was not significant among younger old adults (younger: p = .201; older: p = .001, respectively). In multiple linear regression analysis, Excessive Daytime Sleepiness was associated with slower gait speed among both age groups (younger: p = .021; older: p = .045). Excessive Daytime Sleepiness was not significantly associated with stride length coefficient of variation among young old adults (p = .996), while Excessive Daytime Sleepiness was significant associated with greater stride length coefficient of variation among older old adults (p=.048). |
| Neale | 2022 | Investigate if sleep quantity, quality, and symptoms from self-reported hospital anxiety and depression scores are associated with physical activity. | The fully adjusted rate ratio for steps per day for Time in Bed (hours) [Rate Ratio 0.97(95% CI: 0.92; 1.02)], Nocturnal Sleeping Bouts (numbers) [Rate Ratio 1.02 (95% CI: 0.97; 1.07)] were not significantly associated with number of steps per day, while there was a significantly association with number of steps per day for Hospital Anxiety Scale [Rate Ratio R 1.04 (95% CI: 1.01; 1.07)] and Hospital Depression Scale [Rate Ratio 0.95 (95% CI: 0.91; 0.99)]. |
| O'Dowd | 2017 | Determine if early sleep dysfunction in Parkinson's Disease would predict more rapid deterioration in gait in an incident Parkinson's Disease cohort. | Poor sleep efficiency and greater sleep fragmentation correlated significantly with progression of step-width variability, a gait characteristic mediated by postural control, providing evidence that poor sleep in Parkinson's Disease is associated with a more rapid deterioration in gait. |

| | | | |
|---|---|---|---|
| Oliveira de Almeida | 2021 | Determine if poor sleep quality was associated with Freezing of Gait severity, with all three components of the Freezing of Gait phenotype (cognitive, anxiety, and mobility), and with disease severity; Verify if Freezing of Gait, cognition, anxiety, and mobility explained the variance of the Pittsburgh Sleep Quality Index scores in Parkinson's Disease + Freezing of Gait; Compare sleep quality, cognitive function, anxiety, and mobility among Parkinson's Disease + Freezing of Gait, Parkinson's Disease - Freezing of Gait and Healthy Control. | Significant correlations were observed in the in the Parkinson's Disease + Freezing of Gait group between the Pittsburgh Sleep Quality Index scores and the Timed Up & Go time ($P \leq 0.0038$); In the Parkinson's Disease - Freezing of Gait group, no significant correlations were found between the Pittsburgh Sleep Quality Index scores and the TUG time (r=-0.01, P = 0.962); In the Healthy Controls group, there were significant correlations between the Pittsburgh Sleep Quality Index scores and the TUG time (r = 0.72, P < 0.0001). |
| Overcash | 2018 | Determine the relationship among gait, grip strength, cognition, depression, pain, and fatigue; Identify which variables are most predictive of poor sleep. | Timed Up and Go was not significantly associated with having poor sleep in both unadjusted and adjusted logistic regression analyses. |
| Pan | 2017 | Determine the associations of self-reported sleep quality and duration with health-related quality of life in older Chinese adults. | Poor sleep quality was significantly associated with self-reported problems in mobility, with an Odds Ratio of 2.17 (95% Confidence Interval: 1.15, 4.10); Intermediate sleep quality was related to problems in mobility. (not statistically significant) (Odds Ratio = 1.81, 95% Confidence Interval: 0.99, 3.33); Sleeping shorter than 7 hours was found to be associated with problems with mobility with an Odds Ratio of 4.09 (95% Confidence Interval: 1.58, 10.57). |
| Papazisis | 2021 | Evaluate the correlations between sleep and body mass index in relation to physical activity and food intake during the second lockdown of the COVID-19 pandemic in Greece. | Those with decreasing walking time reported the highest percentage of decreased sleep quality (p = 0.006) and worsened sleep quality (p = 0.016) |
| Park | 2014 | Examine the connection between sleep complaints and incident disability in older adults without disability. | Models controlling for age, sex, and education, a one-point higher dyssomnia score at baseline was associated with about 27% increased risk of mobility disability (hazard ratio: 1.27; 95% Confidence Interval: 1.09-1.48; $x^2_1$=11.04; p <0.01). The association between sleep complaints and incident mobility disability remained significant after controlling for depressive symptoms. |
| Redeker | 2010 | Evaluate insomnia symptoms and the extent to which they are associated with clinical and demographic patient characteristics, daytime symptoms, and functional performance in patients with stable heart failure. | Participants without insomnia symptoms walked approximately 100 ft further (mean 976.8; 95% Confidence Interval 882.9,1070.7) than those with insomnia symptoms (mean 875.3, 95% Confidence Interval 791.0,959.6). |
| Reimers | 2021 | Investigate the impact of physical activity (intensity levels and duration) during the day (7–12 h, 12–18 h, 18–23 h) on sleep quality in patients suffering from idiopathic restless leg syndrome. | Low but significant negative correlation coefficients between the number of steps between 6-11pm and total sleep time (r = -0.2221, 95% Confidence Interval: -0.3827- -0.0432; p=0.0126 (more steps associated with shorter sleep time) and between steps between 6am-12 noon and nighttime Periodic Limb Movements during Sleep (r = -0.1958, 95% Confidence Interval: -0.3768- -0.0003; p = 0.0496; more steps taken in the morning associated with less Periodic Limb Movements during Sleep. |
| Robinson | 2022 | Examine the association between daily physical activity and sleep outcomes across varying levels of insomnia severity. | Days with more steps were associated with nights with longer sleep duration (γ = 0.01, p = 0.004). Daily steps were not significantly associated with better sleep efficiency or quality that night. Baseline insomnia severity was negatively associated with mean daily sleep quality (γ = −0.07, p = 0.026); Insomnia severity was not significantly associated with sleep duration or efficiency; Baseline insomnia severity moderated the relationship between daily steps and sleep duration (γ = −0.001, p = 0.005); There was a significant positive association between daily steps and sleep duration for those with mild insomnia (γ = 0.003, p = 0.022), and a significant negative association between daily steps and daily sleep duration for those who had severe insomnia (γ = −0.004, p = 0.033). |
| Roh | 2014 | Describe the levels of mobility in older cancer patients receiving palliative care in Korea; Examine the associations of their mobility with lifestyle factors (sleep disturbance, physical activity) and physical symptoms (pain, fatigue). | Higher levels of mobility were correlated with lower levels of sleep disturbance (r= -.37). A significant predictor for mobility was levels of sleep disturbance. |
| Rosenbaum | 2016 | Determine whether posttraumatic stress disorder symptom severity and psychological and functional variables were associated with physical activity upon admission to an inpatient facility. | A significant negative association was found between time spent walking and sleep behavior (Pittsburgh Sleep Quality Index) (r = −0.24, p < 0.05; medium effect size, Cohen's d = 0.5), indicating less time walking is associated with poor sleep quality. |
| Saraiva | 2022 | Assess the changes in gait speed and hemodynamics response on the prefrontal cortex resulting from the addition of a cognitive task during walking (cognitive dual-task) compared to normal walking (single task); Determine the correlation between sleep quality and physical activity level with gait performance and brain hemodynamics changes during the dual-task. | There was no significant association between Pittsburgh Sleep Quality Index and gait speed in single task (rho = 0.133, p=0.610) or dual task (rho = 0.301, p=0.214). |

| | | | |
|---|---|---|---|
| Serrano-Checa | 2020 | Analyze the associations of sleep quality, anxiety, and depression with functional mobility, gait speed, and dynamic balance in community-dwelling post-menopausal women $\geq$60 years. | Gait speed (r= -0.248, p<0.001) and Timed Up and Go (r = 0.205, p<0.001) significantly correlated with the Pittsburgh Sleep Quality Index total score; Pittsburgh Sleep Quality Index domains of sleep quality, sleep latency, sleep duration, sleep efficiency, and use of sleeping medication significantly correlated with gait speed; sleep disturbances and daytime dysfunction did not; All Pittsburgh Sleep Quality Index domains except sleep duration and sleep efficiency significantly correlated with Timed Up and Go test; Multivariate linear regression analysis showed poor sleep efficiency (p = 0.000) and the use of use of sleeping medication (p=0.032) was associated with decreased gait speed, and the use of sleeping medication was associated with Timed Up and Go test (p=0.023). |
| Shibata | 2013 | Examine the correlations among objective sleep variables, sleep-wake cycle parameters, and daily physical activity in hemodialysis patients and controls. | The number of steps taken showed only a weak negative correlation to Wake After Sleep Onset (r= -0.308, p < 0.05). No other correlations between steps and sleep. |
| Shih | 2017 | Explore the relationship between sundown syndrome and sleep quality; Determine whether the severity of dementia, sleep quality, and/or weekly duration of walking may influence sundown syndrome; Examine differences in sundown syndrome and sleep quality in relation to the accompanying walker and weekly duration of walking among community-dwelling people with Alzheimer's Disease. | Longer walking time led to better sleep quality ($F_{4,179}$ = 3.592, P= .008). |
| Siengsukon | 2018 | Examine the relationship between sleep quality and cognitive and physical function in individuals with mild multiple sclerosis. | There were no statistically significant difference between the groups on performance of walking ability (6-Minute Walk Test) or common Activities of Daily Living (Physical Performance Test), but the individuals with good sleep quality (Pittsburgh Sleep Quality Index<= 5) had significantly higher scores on the self-report functional abilities (Functional Status Questionnaire) compared to those with poor sleep quality. |
| Silva | 2023 | Investigate the impact of sleep on different domains of physical activity in daily life in idiopathic pulmonary fibrosis. | In the idiopathic pulmonary fibrosis group, sleep duration at night associated significantly with step counts (−0.82 ≤ R ≤ 0.43; p < .05); Compared to controls, the idiopathic pulmonary fibrosis subjects presented lower step counts, less time spent in standing position, and more time spent in lying position (p < .05). |
| Silva | 2021 | Investigate the sleep state and determine whether variables, such as age, functional status, walking capacity, fatigue, depressive symptoms, and quality of life are associated with sleep quality of individuals with chronic stroke. | Significant correlation coefficients were found between sleep quality and walking capacity (r= -0.25; p < .01). In other words, worse sleep quality was associated with worse walking capacity. |
| Spira | 2012 | Determine whether objectively measured sleep quality predicts five-year incident instrumental activities of daily living impairment and decline in grip strength and gait speed in older women. | Women in the shortest total sleep time quartile had double the odds of declining grip strength, compared to those with the longest total sleep time (Adjusted Odds Ratio = 1.97, 95% Confidence Interval 1.17, 3.32); Women in the quartiles with the most wake after sleep onset and the lowest sleep efficiency had an approximately 90% greater odds of grip strength decline than those with the least wake after sleep onset (Adjusted Odds Ratio = 1.90, 95% Confidence Interval 1.11, 3.24) and sleep efficiency (Adjusted Odds Ratio =1.92, 95% Confidence Interval 1.12, 3.29). |
| Stack | 2006 | Describe turning strategies and explore the association between mobility and sleep in patients with Parkinson's Disease. | Using multiple turning strategies to initiate and maintain sleep in those with Parkinson's Disease was associated with self-reported sleep disturbance. |
| Stenholm | 2011 | Characterize elderly persons into sleep/rest groups based on their self-reported habitual total sleeping time and habitual time in bed; Examine the prospective association between sleep/rest behavior on physical function decline. | Both long (≥ 9 h) Total Sleep Time and long Time in Bed predicted accelerated decline in objectively measured physical performance and greater incidence in subjectively assessed mobility disability, but short (≤ 6 h) Total Sleep Time did not. After combining Total Sleep Time and Time In Bed, long sleepers (Total Sleep Time and Time in Bed ≥ 9 h) experienced the greatest decline in physical performance and had the highest risk for incident mobility disability in comparison to mid-range sleepers with 7-8 h Total Sleep Time and Time in Bed. Subjective short sleepers reporting short (≤ 6 h) Total Sleep Time but long (≥ 9 h) Time in Bed showed a greater decline in Short Performance Physical Battery score and had a higher risk of incident mobility disability than true short sleepers with short (≤ 6 h) Total Sleep Time and Time In Bed ≤ 8 hours. |

| | | | |
|---|---|---|---|
| Stenholm | 2010 | Examine whether self-reported sleep duration, insomnia-related symptoms, and fatigue are associated with walking speed and self-reported mobility limitation in men and women aged 55-64 and 65 or more years. | After adjusting for lifestyle factors and diseases, longer sleep (≥9 hours) was associated with a decreased walking speed in women aged 65 or more years (p = .04) and shorter sleep (≤6 hours) with a higher odds for mobility limitation in women aged 65 or more years (Odds ratio = 1.68, 95% Confidence Interval = 1.02–2.75) and in men aged 55–64 years (Odds Ratio = 3.62, 95% Confidence Interval = 1.40–9.37) compared with those having a mid-range sleep duration. Sleeping disorders or insomnia was independently associated with both decreased walking speed and mobility limitation in men aged 55 or more years but only with mobility limitation in women aged 65 or more years. |
| Sullivan Bisson | 2019 | Determine whether a 4-week low-impact physical activity intervention can affect sleep in healthy adults. | Averaged across the month, daily active minutes were positively related to sleep quality, but not duration; Gender moderated this relationship-- women who took more steps and were more active reported sleeping better than those less active; Within-persons, on days that participants were more active than average, they reported better sleep quality and duration in both genders. |
| Suri | 2015 | Examine the association between walking pace and sleep-disordered breathing in the population-level Multi-Ethnic Study of Atherosclerosis. | Slower walking speed was associated with an increased risk of all outcomes indicative of sleep-disordered breathing, including physician-diagnosed sleep apnoea; These risk differences were adjusted for sex, age, and ethnic group; People who walked relatively slowly had 1.5-times the risk of having sleep apnoea compared with people who walked at a faster pace. |
| Takemura | 2021 | Examine the association between subjective and objective measures of sleep parameters and performance-based measures of physical function in advanced lung cancer patients. | Total sleep time was significantly associated with the 6-Minute Walk Test, Timed Up and Go and Sit-to-Stand Test. Pittsburgh Sleep Quality Index global score was only significantly associated with Timed Up and Go (β= 0.140; 95% Confidence Interval = 0.000, 0.280; P= 0.050) after adjustment for multiple covariates; Shorter sleep duration significantly predicted poorer physical performance in advanced lung cancer patients, and more attention is required for those with less than 4.3 hours of sleep on average. |
| Teas | 2021 | Examine the relationship between self-reported and objectively measured sleep and functional capacity in adults. | Better self-reported sleep quality predicted stronger grip, quicker gait, and faster chair stands; Greater Wake After Sleep Onset predicted slower gait speed; Long (>8h) sleep duration and a more variable sleep schedule predicted lower grip strength; Pittsburgh Sleep Quality Index remained a significant predictor of functional measures. |
| Theodorou | 2020 | Investigate the physical activity and quality of sleep, during three days (previous day of dialysis, on the day of dialysis and after day of dialysis), in patients with end-stage renal on hemodialysis. | Differences between three days of average of steps and distance and Pittsburgh Sleep Quality Index parameters 'engaging in social activity' (steps, p = 0:006, distance, p = 0:006) and 'enthusiasm to get things done' (steps, p = 0:029, distance, p = 0:030); Study suggests interrelationship between sleep quality and physical activity. |
| Tighe | 2021 | Investigate the association between better multidimensional sleep health and three performance-based measures of physical functioning: gait speed, lower extremity strength, and grip strength. | Higher levels of sleep health were significantly associated with faster gait speed; Each additional good sleep health domain (regularity of sleep timing, sleep quality, daytime alertness, sleep timing, sleep efficiency, and sleep duration) is associated with a .03 meter per second faster gait speed. |
| Tyagi | 2015 | Examine the association between self-reported daytime sleepiness, mobility, and balance in community-dwelling elderly adults. | Participants with daytime sleepiness walked slower than those without (1.01 +/- 0.25 vs 1.13 +/- 0.24 m/s; P=.01), and the difference persisted after controlling for covariates (adjusted difference 0.09 +/- 0.04; P=.03). |
| Umemura | 2022 | Explore the association between walking test performance, the cognitive status, and the parameters of circadian expression in stroke survivors. | Subjects who perform better in the walking tasks have higher daily activity levels and a more stable circadian rhythm, with lower daily fragmentation and a more stable synchronization with the 24-hour light/dark cycle. |
| Umemura | 2021 | Identify changes in gait performance associated to sleep disturbances. | There was a statistically significant difference (p<0.05) between the Control Group and Sleep Chronic Restriction groups in the frequency early and late footfalls; Accuracy was less for Sleep Chronic Restriction in early but better in late. No differences were observed between these groups for the hit interval (− 30 ms <RP> 10 ms). Acute sleep deprivation (one night) group exhibited impaired performance in the sensorimotor synchronization gait protocol, such as a decrease in the Period Error between the footfalls and the auditory stimulus & missed more frequently the auditory cues. The group with chronic sleep restriction also underperformed when compared to the control group with a tendency to a late footfall with respect to the Rhythmic Auditory Cueing sound. |
| Vardar-Yagli | 2015 | Assess the impact of sleep quality on functional capacity, peripheral muscle strength, and quality of life in patients with Chronic Obstructive Pulmonary Disease. | Pittsburgh Sleep Quality Index total score was significantly correlated with physical mobility; The Pittsburgh Sleep Quality Index sleep latency score showed a negative correlation with 6-Minute Walk Test distance; There was a positive correlation between Pittsburgh Sleep Quality Index daytime dysfunction score and 6-Minute Walk Test distance; Pittsburgh Sleep Quality Index sleep latency is independently predicted by 6-Minute Walk Test distance. |
| Vaz Fragoso | 2014 | Evaluate sleep-wake disturbances in sedentary community-dwelling elders with functional limitations. | Participants with insomnia, daytime drowsiness, and poor sleep quality had mean values of 12.1 for Insomnia Severity Index, 12.5 for Epworth Sleepiness Scale, and 9.2 for Pittsburgh Sleep Quality Index, respectively; In adjusted models, measures of mobility and physical inactivity were generally not associated with sleep-wake disturbances, using continuous or categorical variables. |
| Wang | 2022 | Investigate the association between sleep quality, sleep duration , and gait speed among Chinese older adults. | In the adjusted model, poor sleep quality and longer sleep duration were significantly associated with slower normal walking speed in Chinese adults (p < 0.001). Moreover, there were negatively significant associations between normal gait speed and sleep quality in Male adults (p < 0.01). |

| | | | |
|---|---|---|---|
| Xue | 2018 | Evaluate the relationships between symptoms of nocturnal hypokinesia and sleep quality in Parkinson's Disease patients by utilizing multisite inertial sensors and polysomnography. | Subjective sleep quality scores, including the Pittsburgh Sleep Quality Index (P = 0.011) and Parkinson's Disease Sleep Scale (P = 0.009) were significantly different between these (Parkinson's Disease + Impaired Bed Mobility vs Parkinson's Disease - Impaired Bed Mobility) two groups; Parkinson's Disease + Impaired Bed Mobility group had less TST (P = 0.006) than the Parkinson's Disease - Impaired Bed Mobility group; Total Sleep Time showed positive correlation with the number of turning-over events (r = 0.505, P < 0.05); Sleep Efficiency was positively correlated with the number of turning-over events (r = 0.473). |
| Yeom | 2015 | Describe the levels of mobility in community-dwelling older Koreans with chronic illnesses; Examine the associations of mobility with lifestyle factors (sleep patterns, physical activity) and physical symptoms (fatigue, pain). | Mobility was significantly associated with participants' perceptions of the quality of their sleep depth and duration; Participants who rated the quality of their sleep as deep showed a higher level of mobility than those reporting their depth of sleep as light (t= -2.44, p= .015). Participants who perceived their sleep duration as long also reported a higher level of mobility than persons who perceived their sleep duration as short (t= -2.79, p= .006). |
| Zhan | 2023 | Examine correlation between sleep duration and physical performance in patients undergoing hemodialysis. | Timed Up and Go in group <7 h and ≥9 h was higher than mid-range sleep duration; Gait speed in group <7 h and ≥9 h was lower than group 7- h; Slower walk speed was associated with <7 h and ≥9 h sleep duration. |
| Zhang | 2021 | Examine the gender-specific associations of sleep quality with gait speed and falls among older adults; Explore the possible mediating effect of muscle strength on these relationships. | In men, higher sleep quality was positively correlated with gait speed (β = 0.008, p = 0.031); In women, higher sleep quality was also positively correlated with gait speed (β = 0.008, p = 0.017). Grip strength mediated these associations in men but not in women, and the mediating effects of grip strength can explain 23.74% of the total effect of sleep quality on gait speed. |
